# Changes in CD163+, CD11b+, and CCR2+ peripheral monocytes relate to Parkinson’s disease and cognition

**DOI:** 10.1101/2021.03.15.21253572

**Authors:** Sara Konstantin Nissen, Kristine Farmen, Mikkel Carstensen, Claudia Schulte, David Goldeck, Kathrin Brockmann, Marina Romero-Ramos

**Affiliations:** DANDRITE & Department of Biomedicine, Aarhus University, Aarhus, Denmark; Center of Neurology, Department of Neurodegeneration and Hertie-Institute for Clinical Brain Research & German Center for Neurodegenerative Diseases, University of Tuebingen, Tuebingen, Germany; Department of Internal Medicine II, Centre for Medical Research, University of Tuebingen, Tuebingen, Germany

**Author notes:** **Corresponding author:** Marina Romero-Ramos, Dept. of Biomedicine, Aarhus University, Høegh-Guldbergsgade 10, DK-8000 Aarhus C, Denmark.

**Keywords:** Alpha-synuclein, neuroinflammation, dendritic cells, natural killer cells, TLR, monocytes, Parkinson’s disease

## Abstract

1.

Alpha-synuclein pathology is associated with immune activation and neurodegeneration in Parkinson’s disease. The immune activation involves not only microglia but also peripheral immune cells, such as mononuclear phagocytes found in blood and infiltrated in the brain. Understanding peripheral immune involvement is essential for developing immunomodulatory treatment. Therefore, we aimed to study circulating mononuclear phagocytes in early- and late-stage Parkinson’s disease, defined by disease duration of less or more than five years, respectively, and analyze their association with clinical phenotypes. We performed a cross-sectional multi-color flow cytometry study on 78 sex-balanced individuals with sporadic Parkinson’s disease, 28 controls, and longitudinal samples from seven patients and one control. Cell frequencies and surface marker expressions on natural killer cells, monocyte subtypes, and dendritic cells were compared between groups and correlated with standardized clinical scores. We found elevated frequencies and surface levels of migration-(CCR2, CD11b) and phagocytic-(CD163) markers, particularly on classical and intermediate monocytes in early Parkinson’s disease. HLA-DR expression was increased in advanced stages of the disease, whereas TLR4 expression was decreased in women with Parkinson’s Disease. The disease-associated immune changes on CCR2 and CD11b correlated with worse cognition. Increased TLR2 expression was related to worse motor symptoms. In conclusion, our data highlights the TLR2 relevance in the symptomatic motor presentation of the disease and a role for peripheral CD163^+^ and migration-competent monocytes in Parkinson’s disease cognitive defects. Our study suggests that the peripheral immune system is dynamically altered in Parkinson’s disease stages and directly related to both symptoms and the sex bias of the disease.

**Highlights:**

- TLR2 expression increased in patients with worse motor symptoms.
- Increased CD163 and HLA-DR monocytic expression in patients with long PD duration.
- Sexual-dimorphism for CCR2, CD11b, and TLR4 expression on PD monocytes.
- CCR2 and CD11b expression are associated with cognitive impairment in PD.

## 2. Introduction

The immune component of Parkinson’s disease (PD) has been settled by studies showing microglia activation in *post-mortem* brains (Imamura et al., 2003b), low-level systemic inflammation (Qin et al., 2016), and decreased PD risk in NSAID users (Gagne and Power, 2010; Ren et al., 2018). Moreover, variants in immune-related genes, including HLA-DR, are associated with increased PD risk (International Parkinson Disease Genomics et al., 2011; Rayaprolu et al., 2013). This suggests a significant role for both the innate and the adaptive immune systems. Accordingly, both CD8 and CD4 T-cells have been associated with the disease. Decreased number of T-cells in blood from people with PD (PwP) relates to disease severity and seems to parallel infiltration of CD8 and CD4 T-cells into the brain (Bhatia et al., 2021; Galiano-Landeira et al., 2020; Kouli et al., 2020). Indeed, IFN-gamma producing cytotoxic T-cells are elevated in the blood of PwP (Yan et al., 2021) and infiltrate the midbrain prior to the dopaminergic neuronal death (Galiano-Landeira et al., 2020). Additionally, T-cells from PwP exhibit a Th1/Th17 bias (Storelli et al., 2019), with T-cells responding to α-synuclein (α-syn)-derived peptides early in PD (Lindestam Arlehamn et al., 2020; Sulzer et al., 2017). Despite the importance of the innate immune system’s role in activating/priming the adaptive system, there is a paucity of evidence on the innate peripheral immune cells’ changes in PD. Most research has focused on microglia; however, peripheral immune cells can infiltrate the brain and participate in the inflammation and neurodegeneration in PD (Harms et al., 2021). Moreover, increasing evidence suggests that PD is not only a CNS disorder but also affects and, in some cases, originates in the periphery (Horsager et al., 2020). For these reasons, more knowledge regarding the peripheral innate immune cells is required.

We have previously shown that biofluids from PwP had increased levels of soluble (s)CD163, a marker produced upon activation of CD163-expressing mononuclear phagocytes (MNPs) (Nissen et al., 2021). This elevation was related to PD duration(-stages), and associated with cognition and neuronal disease biomarkers such as α-syn, Tau, and phosphorylated-Tau. This suggests an active involvement of MNPs in the different stages of PD. MNPs comprise populations of ontogenic, phenotypic, and functionally overlapping monocytes (Mos) and dendritic cells (DCs) (Dutertre et al., 2019; Geissmann et al., 2003). MNPs act as myeloid precursors that enter the tissue and differentiate into Mo-derived macrophages or DCs, (Dutertre et al., 2019; Geissmann et al., 2003). Daily, half of the circulating MNPs leave the bloodstream under steady-state conditions to infiltrate tissue, which can increase during inflammation (Geissmann et al., 2003). Based on their expression of CD14 and CD16, Mos are typically subdivided into classical (cMo, CD14^high^/CD16^-^), intermediate (iMo, CD14^low/high^/CD16^+^), and non-classical (ncMo, CD14^-/low^/CD16^++^) (Abeles et al., 2012). cMos, and to some degree also iMos, migrate to inflamed tissue in a CCR2-dependent manner and are highly phagocytic. In contrast, ncMos are described as blood-endothelia patrolling cells migrating to non-inflamed tissue, including the brain, in a CX_3_CR1-dependent manner (Geissmann et al., 2003). MNPs also include a fraction of the dendritic cells (CD14^-/low^/CD16^-^), responsible for antigen presentation to and priming of T-cells. Although DCs, in general, do not express CD14 and CD16, many of them share many other surface receptors with Mos, such as TLR2, HLA-DR, CCR2, and CD163. Studies in PD animal models suggest infiltration of CCR2^+^ or CD163^+^ cells partaking in the inflammatory and neurodegenerative processes associated with PD (Harms et al., 2018; Tentillier et al., 2016).

Immune changes in the brain (Edison et al., 2013) and changes on MNPs are suggested to be highly relevant in PwP with significant cognitive decline (Nissen et al., 2021; Wijeyekoon et al., 2020a). Interestingly, the presentation of PD differs between sexes (Iwaki et al., 2021), with cognitive impairment being less likely to occur in female patients. We have previously reported a sex dimorphism in the CD163 related immune changes in serum from PwP (Nissen et al., 2021). In addition, transcriptomic analysis of blood CD14^+^ Mos has recently shown that females with PD show a homogenous monocytic response, while the response was more heterogeneous in males (Carlisle et al., 2021). All this supports a sex-specific immune response that might be associated with the differences in disease presentation, especially regarding cognition.

To study the profile of peripheral MNPs *ex vivo* in PD, we used multicolor flow cytometry on a cross-sectional cohort of patients with sporadic PD and seven longitudinal PD cases. Based on our prior data, we divided the patients in the cohort into two groups: those whose samples were collected early after disease diagnose (early PD) and those collected late in the disease (late PD) using five years as a cut-off. We analyzed putative sex differences and then correlated the data to the motor and non-motor symptoms.

## 3. Materials and methods

### 3.1 Cohorts and sample collection

All participants were examined by a neurologist specialized in movement disorders. PD was diagnosed according to UK Brain Bank Society Criteria (Litvan et al., 2003). PD patients were assessed in the dopaminergic ON state. We evaluated the severity of motor symptoms using part III of the Unified Parkinson’s disease Rating Scale (UPDRS-III), from 2006-2008 the old version and from 2009 onwards the MDS-UPDRS (Goetz et al., 2008). The disease stage was categorized by the modified Hoehn and Yahr Scale (H&Y) (Goetz et al., 2004). Cognitive function was tested using the Montreal Cognitive Assessment (MoCA) (Hoops et al., 2009) and/or the Mini Mental Status Examination (MMSE) (Folstein et al., 1975). Since the MoCA was available only from 2009, all previously obtained MMSE scores were converted into MoCA equivalent scores according to published algorithm (Bergeron et al., 2017). Cognitive impairment was defined if the MoCA score was ≤26 based on Hoops et al. (point of maximum sensitivity without losing specificity) (Hoops et al., 2009). Mood disturbances were assessed using Beck’s Depression Inventory II (BDI II) (Beck et al., 1996). Scores above 13 were considered as “depressed mood” (Kuhner et al., 2007). Olfactory dysfunction was detected using the *Sniffin’ Sticks*. A cutoff of <9 (<75%) of 12 odors indicated hyposmia (Hummel et al., 2007). Subjects with respiratory allergies or infections were excluded from this test. Levodopa-equivalent-dosage was calculated according to the German Society for Neurology (http://www.dgn.org/). Patients’ spouses and volunteers served as control participants after being medically assessed to have no neurodegenerative disease (**Supp.Fig.1**). The local ethics committee approved the study (480/2015BO2), with all participants providing written informed consent. Peripheral blood mononuclear cells (PBMCs) were collected at the date of the respective clinical visit when scores were assessed, isolated, and stored at the Hertie Neuro-Biobank (University of Tuebingen) and analyzed at Aarhus University (see supplementary information). The study includes *Cohort#1A:* Cross-sectional samples from 78 PwP collected early (< 5 years) or late (≥5 years) after PD diagnosis and 28 HCs (**Table 1**); and *Cohort#1B*: Longitudinal samples from seven PwP and one HC (**Supp.Table 1**). Information on comorbidities and pharmacological treatment for *Cohort#1A* is provided in **Supp.Table 2**. From the samples received (n=109), three individuals with known autoimmune comorbidities were excluded: one female control and two late PD, one female and one male (**Supp. Fig.1**).

**Table 1:**
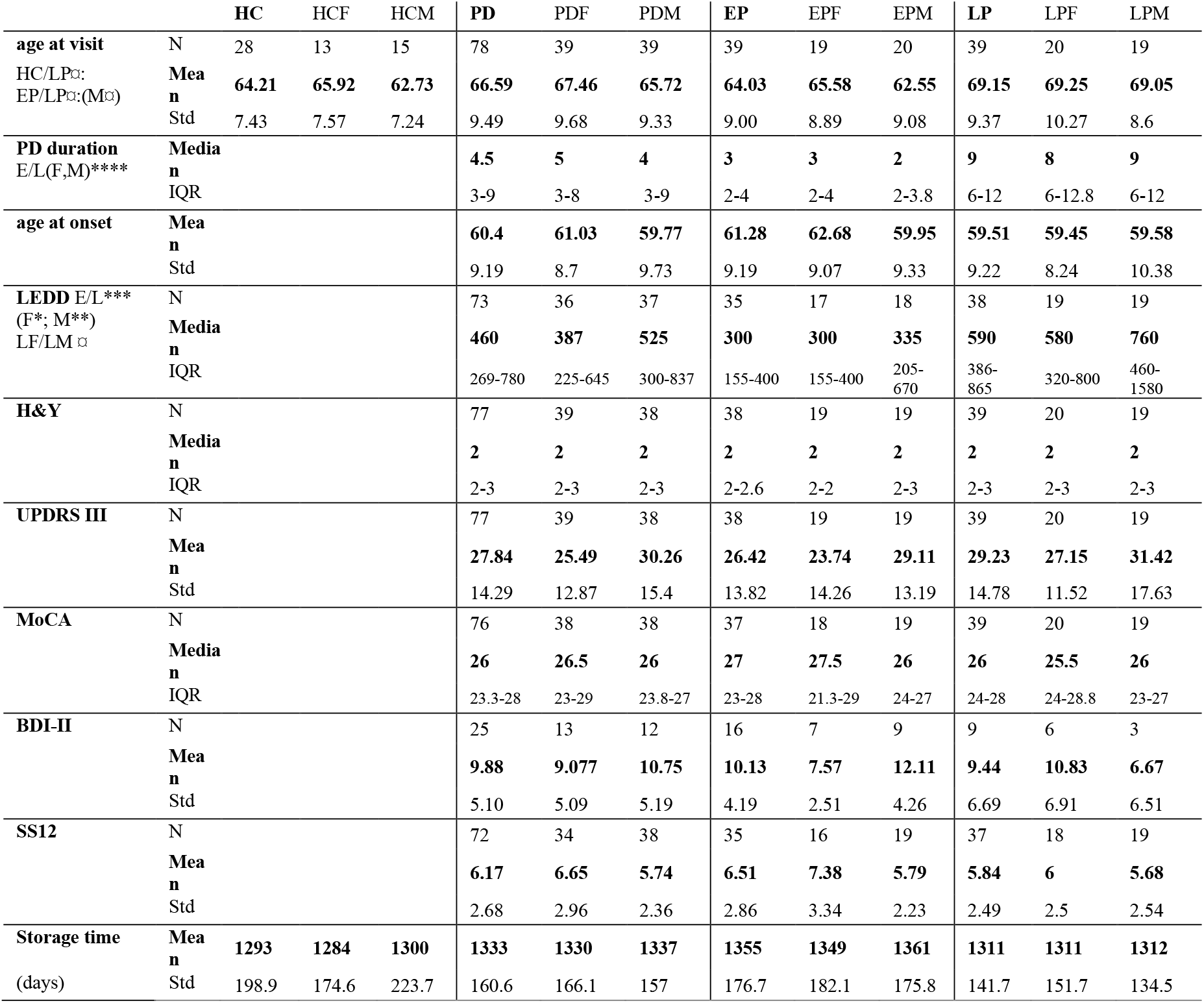
Clinical information for the cross-sectional *Cohort#1A*. *Cohort#1A* for cross-sectional analysis of blood samples collected from healthy control (HC) and people with sporadic Parkinson’s disease (PD) early (<5 years, EPD) or late (≥5 years, LPD) after diagnosis, without known autoimmune comorbidities, and with an equal distribution between females (F) and males (M). Mean, standard derivation (SD) for normally distributed data, median with interquartile range (IQR) for non-parametric data, and the number (N) of individuals for whom the data were available from at the sampling date is shown for: L-dopa equivalent daily dose (LEDD), Hoehn and Yahr (H&Y), Unified Parkinson’s Disease Rating Scale three (UPDRS III), the Montreal Cognitive Assessment (MoCA) score, Beck Depression Inventory II (BDI II), and Sniffin’ Stick 12 olfaction test (SS12). ¤/* parametric and non-parametric significant p values are shown the corresponding unpaired t-tests comparison or one-way ANOVA * p<0.05, ** p<0.01, *** p>0.001, **** p<0.0001.

### 3.2 Flow cytometry

PBMCs were stained for flow cytometry with a combination of nine fluorochrome-conjugated antibodies (**Supp.Table 2**), as previously described (Farmen et al., 2021). The cells were run on an LSR Fortessa (BD) flow cytometer, and data were analyzed using FlowJO V10. For details, see supplementary information.

### 3.3 Statistical analyses

GraphPad Prism V7 and JMP (V14) were used for statistical analyses. In *Cohort#1A*, normality was informed with D’Agostino-Pearson omnibus test or residual QQ plots. Differences in clinical and demographic data were tested by parametric or non-parametric unpaired t-test and one-way ANOVA with Fishers uncorrected LSD. Outliers were identified using ROUT (Q=0.1%). On gaussian distributed raw flow cytometry data or log-transformed data, sex, disease group, and sex-disease group interactions were analyzed using two-way ANOVA. When sex influenced the outcome, sexes were separated and groups within one sex compared with Bonferroni correction for multiple comparisons. If sex did not affect the result, the interaction was removed from the analysis, and the disease groups were compared using a post hoc Tukey’s multiple comparison test, and the sexes merged in the graphs. Associations between flow cytometry measurements and clinical data were tested using Pearson (or Spearman) correlations as appropriate with two-tailed p values, and if significant, assessed for the relevant covariates (age-at-visit and levodopa equivalent daily dosage (LEDD)) using simple and multiple linear regressions. Storage time for the frozen samples in each group was similar, hence not used as a covariate. Only associations with p<0.05 for correlation and linear regression were considered unless indicated. Assessment of differences related to clinical thresholds was compared with parametric or non-parametric unpaired t-tests, as relevant.

For comparison of longitudinal in *Cohort#1B*, normality was tested on residual QQ plots, with no apparent deviations from the reference line that could reject normality. To compare sampling at different months, paired repeated-measures mixed-effect model with Tukey’s multiple comparisons were performed. The first and last visits were compared with parametric paired t-tests.

We evaluated the effect of anti-inflammatory drugs and found no difference in the mean between groups of individuals with such treatment and without. Furthermore, the values of these individuals were evenly spread. Therefore, they were included in the analysis.

## 4. Results

### 4.1 Demographics

After autoimmune comorbidity exclusion, *Cohort#1A* included sporadic 78 PD patients and 28 HCs with similar sex-(50%) and age-distribution (HC 64.2±7.4 & PD 66.6±9.5; mean±SD). We subdivided the PD patients into two groups according to the disease duration at sampling: early (<5y) or late PD (≥5 years) based on our prior data on soluble (s)CD163, which supported a stage-associated (disease duration) increased MNP activation in PwP (Nissen et al., 2021). Equally, that data also indicated a sex difference in the MNP activation in patients’ blood; thus, sex differences were interrogated for each analysis (see methods). This subdivision resulted in a late PD group being older than the early PD group and HCs, which was also significant when investigating the males only, but not females (**Table 1**). As expected for disease duration, LEDD was higher in late vs. early PD and higher in late males vs. late females (**Table 1**). Thus, all differences found among these groups were appropriately compensated for the relevant covariates: age and LEDD.

Additional eight individuals provided longitudinal samples: *Cohort#1B*. Two to four samples were available per individual, with a timeframe of one to sixteen months from the first to the last visit (**Supp.Table 1**). *Cohort#1B* was analyzed separately.

### 4.2 Unraveling the MNPs in the PBMC pool by TLR2 expression

Classical gating of Mos by forward- and side scatter before CD14/CD16 subtyping results in an ignored fraction, including circulating DCs (Geissmann et al., 2003). Instead, we used CD56 and TLR2 as inclusion markers for natural killer (NK) cells and MNPs, respectively. MNP (TLR2^+^) included all Mos and some DCs, as shown by t-distributed stochastic neighbor embedding plot of concatenated samples of live cells (**Supp.Fig.2**): All CD14^+^ and CD16^+^, except the CD16^+^/CD56^+^ cells, were TLR2^+^. Hence, our gating distinguishes the (TLR2^+^) MNPs from the (TLR2^-^/CD56^dim^) mature- and (TLR2^-^/CD56^bright^) precursor NK cells from the remaining PBMC pool (TLR2^-^/CD56^-^) (**Supp.Fig.3**).

The TLR2^+^ MNPs were divided into four subpopulations based on CD14/CD16 and HLA-DR expression (other characteristics in brackets): cMos CD14^++^/CD16^-^/HLA-DR^dim^ (CD11b^+^/CCR2^bright^/CD163^bright^/TLR2^bright^), iMos CD14^++^/CD16^+^/HLA-DR^bright^ (/CD11b^+^/CCR2^bright^/CD163^bright^/TLR2^bright^), ncMos CD14^dim^/CD16^++^/HLA-DR^dim^ (/CD11b^-/dim^/CCR2^+^/CD163^dim/-^/TLR2^bright^), and the DCs CD14^dim/-^/CD16^-^/HLA-DR^bright^ (/CD11b^-^ /CCR2^+^/CD163^Mixed^/TLR2^dim^). MFI for the different proteins were only further analyzed in the full MNP population, if the expression appeared homogeneous, or in populations with a clearly defined bright subset (**Supp.Fig.2&3** for full info).

In-depth analysis of DCs discriminates the plasmacytoid (p)DC and the antigen-presenting conventional (c)DC type 1 and 2. An additional antibody panel on three control donors (not included in the *Cohort#1A* nor *#1B*) confirmed that the CD1c^+^ cDC2s are TLR2^+^, thus, included in our MNP population, whereas both the CD141^+^ cDC1s and the CD303^+^ pDCs (Dutertre et al., 2019) were TLR2^-^ (**Supp.Fig.4**). The cDC2 is responsible for Th1/Th17 CD4 T-cell priming (Dutertre et al., 2019). We showed that the TLR2^+^ cDC2s primarily were located in the DC gate being CD14^-/dim^/CD16^-^, with a smaller CD14^++^/CD16^-^ fraction, thus in the cMos gate (**Supp.Fig.4**). With our gating, cDC2s constituted ∼5% of the MNPs, with ∼90% of the CD14^-^/CD16^-^ MNPs being cDC2s. For simplicity, the TLR2^+^/CD14^low^/CD16^-^ are referred onward as DCs.

The percentage of viable cells in the flow staining was increased in late PD (with one outlier removed), although this did not remain significant after adjusting for age (p=0.052) (alone and with LEDD) (**Supp.Fig.5A**). Viability was not associated with freezer storage time. Nevertheless, PBMC viability correlated negatively with the Montreal Cognitive Assessment (MoCA) scores for all PwP, even when adjusted for age and LEDD (**Supp.Fig.5B**). Hence, worse cognition was associated with better PBMC viability. Accordingly, when dividing the cohort according to MoCA score (>26=no cognitive impairment and ≤26=cognitive impairment), the cognitive impaired group had greater cell viability (**Supp.Fig.5C**).

### 4.3 Mature NK cells in PwP are activated and primed for antibody-dependent cytotoxicity

Peripheral NK cells migrate to the CNS during synucleinopathies (Earls et al., 2020). Based on CD56 expression, we separated NK cells into precursor and mature populations. Since CD16 increase on NK cells has been related to activation of NK cells (Earls and Lee, 2020b), we also analyzed the activated CD16^+^ fraction within the mature NK population (**Fig.1A-B**). We observed an increase in CD16 median fluorescence intensity (MFI), i.e., upregulation of the antibody receptor FCγRIII expression, on the mature NK cells during early PD in both sexes. Still, in males, this increased further in late PD (**Fig.1B**). These findings remained after compensating for age (the significance was lost for early males when also adjusting for LEDD).

**Figure 1:**
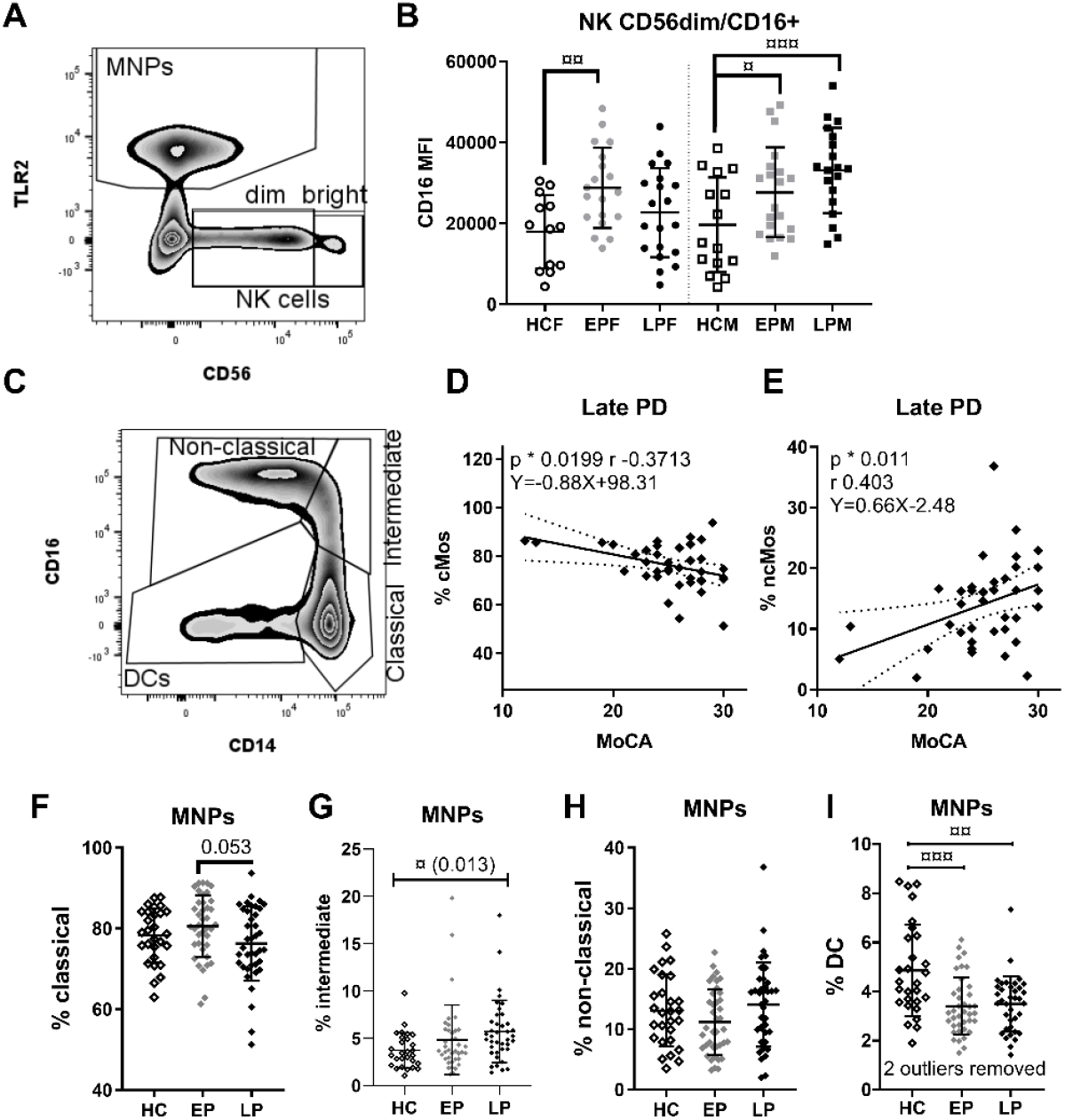
Increased NK cell activation in PwP and changes in MNP subtype distribution during PD. **A)** Gate for identification of all mononuclear phagocytes (MNPs) and natural killer (NK) cells divided into mature (CD56^dim^) and precursor (CD56^bright^) populations. **B)** CD16 median fluorescence intensity (MFI) on CD16^+^ cells within the CD56^dim^ mature NK population in healthy controls (HC), early PD (EP), and late PD (LP) in female (F) and male (M) groups analyzed by Two-way ANOVA with Bonferroni correction for multiple comparisons. **C)** Gating of monocyte subtypes. Plots for the late PD group Spearman correlations (*, p & r) and linear regressions (equation) between Montreal Cognitive Assessment (MoCA) score and the frequency of **D**) classical (cMos) and **E**) non-classical (ncMos) monocytes. Distribution of **E)** cMo, **F)** intermediate (iMo), **G)** ncMo, and **H)** dendritic cells (DCs) (two outliers removed) of the total MNP population. Statistical approach **for F-I)**: Two-way ANOVA (sex/disease group, without interaction) with Tukey’s multiple comparison p values as ¤. **F-I** are interrelated based on the gating; thus, the p values were further Bonferroni corrected for multiple comparisons: p values for DCs, but not iMos (uncorrected shown in bracket) remained below the adjusted threshold of 0.0125. Non-parametric/parametric test: */¤ p <0.05, **/¤¤ p <0.01, ***/¤¤¤ p <0.001.

### 4.4 PD stage-related changes in the distribution of the MNP subtypes

The MNP percentage among PBMCs was similar for HCs (mean±SD: 23.25%±6.60), early (27.01%±8.68), and late PD (24.22%±10.53) and was not affected by stage or sex. However, the MNP subtypes were differently associated with MoCA (see below), and the distribution was altered by the disease stage (**Fig.1C-I)**. Two-way ANOVA with Tukey’s multiple comparisons revealed no sex difference for the MNP subtypes; thus, the data were analyzed without sex-disease group interaction and sexes merged in the graph. The data showed a trending decrease of the cMos from early to late PD (**Fig. 1F**). No changes in the ncMos were seen (**Fig.1H**). However, iMos were significantly increased in late PD vs. HC (**Fig.1G**), with DCs being significantly decreased both in early and late PD (**Fig.1I**). With the different MNP subtypes gating and frequencies being inter-dependent, a Bonferroni correction (alfa 1.25%) was considered, and only the DCs remained significant after this. Notably, we observe a significant, although weak, positive correlation between the percentages of DCs with the Sniffin’ sticks 12 olfaction scores in PwP (**Supp.Fig.5D**). Hence, fewer DCs were associated with hyposmia, not driven by covariates (age and LEDD). Furthermore, in the late PD cases, the fraction of cMos and ncMos correlated negatively and positively with MoCA, respectively (**Fig.1D-E**). Hence, those patients where the number of cMos in blood did not decrease back to healthy levels at late PD (and vice versa with ncMos) were those developing cognitive impairment.

### 4.5 Altered expression of receptors responsible for *α*-syn sensing and presentation

Variants in the gene encoding for the antigen-presenting protein HLA-DR are associated with PD, and fibrillar α-syn increases antigen-presenting capacity on myeloid cells (Harms et al., 2021); hence we investigated its expression in the cohort. The Mo populations expressing a high level of HLA-DR, the iMos and ncMos, showed increased HLA-DR expression (MFI) in PwP (HC vs. PD, iMos p=0.01 and ncMos p=0.013, non-parametric t-tests). Although when separated in stages, this was only significant at late PD (**Fig.2A-B**). The late increase remained significant when adjusting for age. However, these late increases correlated with LEDD, and the significance was lost when compensated for L-DOPA dose.

**Figure 2:**
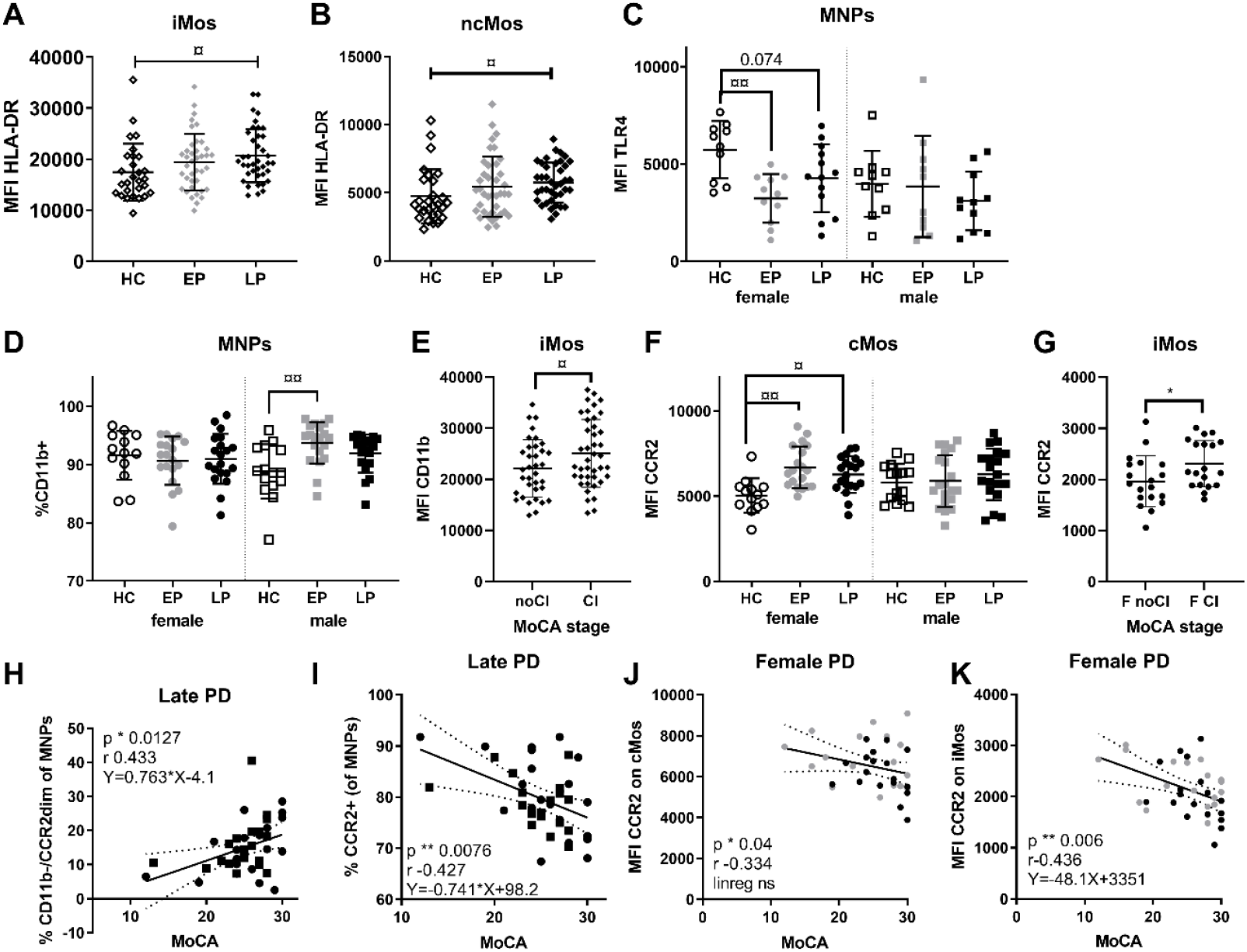
Mononuclear phagocyte expressions and frequencies of HLA-DR, TLR4, CD11b, and CCR2 are changed in PD and associated with cognitive symptoms. Median fluorescence intensity (MFI) of HLA-DR on **A)** intermediate (iMos) and **B)** non-classical monocytes (ncMos) in healthy control (HC), early PD (EP), and late PD (LP) groups. **C)** MFI of TLR4 on all mononuclear phagocytes (MNPs); and **D)** the percentage of CD11b^+^ MNPs, analyzed sex separated. **E**) The MFI of CD11b on iMos in patients with no cognitive impairment (noCI), i.e., Montreal Cognitive Assessment (MoCA) scores >26; or with cognitive impairment (CI, MoCA≤26). **F)** MFI of CCR2 on cMos analyzed by group and sex; **G)** and CCR2 MFI on iMos in females (F) with or without cognitive impairment (CI vs. noCI). MoCA scores in patients in the late PD group correlated **H**) positively (p=0.041 after correction for age covariate with the fraction of CD11b^-^/CCR2^dim^ MNPs (identified in **Supp.Fig.3F**); and **I)** negatively with the total fraction of CCR2^+^ MNPs, (one outlier removed using Grubs, p=0.035 after correction for age covariate). In all females with PD, MoCA was negatively correlated with MFI of CCR2 on **J)** classical monocytes (cMos) and **K)** iMos, but only iMos also showed a significant linear regression that remained after adjusting for age (p=0.03). Statistics: Two-way ANOVA tests with appropriate multiple comparisons. Mean with SD, Spearman correlation p and r values, and linear regression equation with 95% confidence intervals are shown. Uncorrected p values are shown. Non-parametric/parametric test: */¤ p <0.05, **/¤¤ p <0.01, ***/¤¤¤ p <0.001.

We analyzed the innate sensor TLR4, which can recognize α-syn and facilitate α-syn clearance (Harms et al., 2021). Our TLR4 analyses only included 66 samples due to antibody lot number-associated variance. TLR4 expression differed among sexes, and a sex-group interaction was observed; thus, sexes were separated (**Fig.2C**). This revealed a reduced TLR4 expression on early PD females’ MNPs that remained after covariate adjustment (age and LEDD). Thus, HC females had TLR4 levels higher than PD females and all males.

### 4.6 Sex-dependent changes related to MNP migratory capacity

CD11b and CCR2 are molecules important for MNP’s migration and previously related to α-syn (Harms et al., 2021). Therefore, we examined the integrin CD11b (**Fig.2D-E**) and the chemokine receptor CCR2 (**Fig.2F-G**) in MNPs, with a particular focus on cMo and iMo, the cell types showing the highest expression levels for these markers (**Supp.Fig.3**). Although the percentage of CD11b^+^ MNPs was similar among sexes in HCs, we observed an increased percentage of CD11b^+^ MNPs in early PD males vs. HC, but not in females (**Fig.2D**). This remained after covariates adjustment (age and LEDD). We did not find significant changes in the CD11b expression in blood among these groups. However, examination of the MFI CD11b on iMos within all samples from PwP revealed higher CD11b expression in patients with cognitive impairment vs. those without any cognitive impairment, according to MoCA scores (**Fig.2E**)

Regarding CCR2, we observed increased expression on cMos in female patients (**Fig.2F**) that remained after compensating for age and LEDD, while no significant differences were seen in males. The iMos also showed CCR2 upregulation in female patients with cognitive impairment vs. those without (**Fig.2G**). This suggests mobilization of the Mos to inflamed tissue, associated with CD11b in male patients and CCR2 in females, which seems relevant for their cognitive capacity.

Further supporting this, in late PD, the percentage of MNPs with no/low expression of CD11b/CCR2, hence less transmigration capacity, correlated positively with better cognition score in both sexes. Conversely, the percentage of CCR2^bright^ MNPs correlated negatively with cognition (**Fig.2H-I**; both remained after adjusting for age, but the first missed significance after also adjusting for LEDD p=0.062). Furthermore, in females only, the expression of CCR2 on cMos and iMos also had a negative correlation with MoCA (**Fig.2J-K**); for the iMos the linear regression was also significant even when age- and LEDD-adjusted. Overall, changes of markers related to Mo migration were associated with cognitive symptoms, with higher migratory capacity correlated to worse cognition.

### 4.7 Peripheral CD163 changes distinguish PwP from HCs

CD163 is an MNP specific protein whose cleavage from the membrane surface is associated with Mo activation and production of sCD163 (Moller, 2012). We have previously shown that PwP’s blood immune cells *in vitro* have reduced ability to downregulate CD163 expression on CD14^+^ Mos upon exposure to α-syn (Nissen et al., 2019); and that PwP have increased sCD163 levels in serum (females only) and CSF (both sexes), with CSF sCD163 levels associated with α-syn and cognitive deficits (Nissen et al., 2021). This data suggested a role for CD163^+^ cells in PD. Thus, we analyzed the CD163 expression on MNPs. Interestingly, PwP showed higher CD163 expression (MFI) on the high expressing cMo and iMo (bright) compartments (**Fig.3A-B**) that passed covariate testing (age and LEDD). Furthermore, we observed an increased frequency of CD163^+^ MNPs in early PD (**Fig.3D**) that remained after adjusting for age and LEDD.

**Figure 3:**
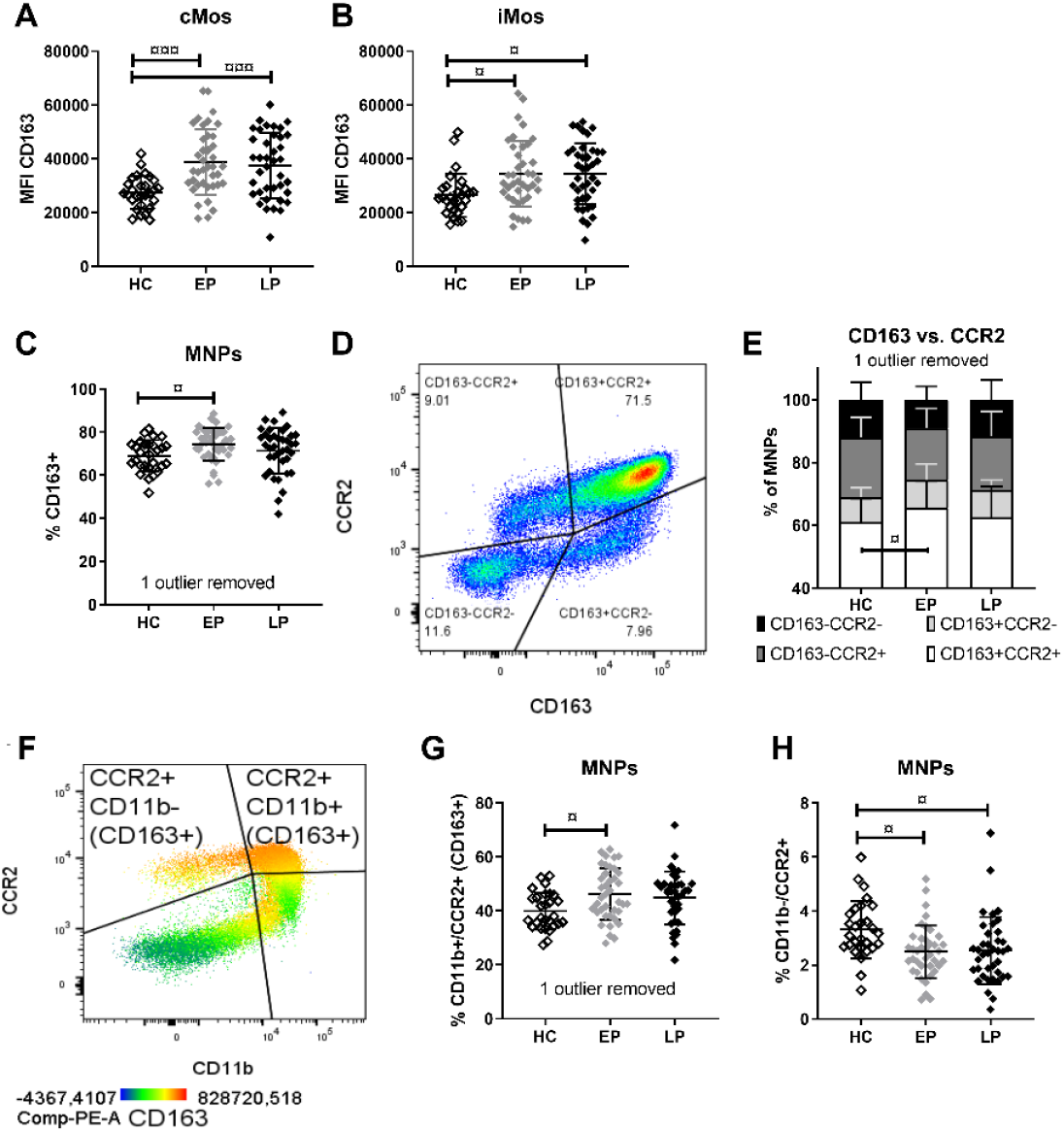
The frequency of CD163^+^ cells and CD163 expression are increased in PwP and related to migration markers. Median fluorescents intensity (MFI) of CD163 on **A)** classical (cMos) and **B)** intermediate (iMos) monocytes. **C)** Frequency of CD163^+^ mononuclear phagocytes (MNPs). **D)** MNPs were gated according to their CD163 and CCR2 expression. **E)** The distribution of the four subpopulations in the groups with shared/depended gating was compared with 2-way ANOVA (disease stage, cell population, interaction) followed by Tukey’s multiple comparison test. **F)** MNPs analyzed based on CCR2 and CD11b, with the majority of CCR2^+^ MNPs, being also CD163^+^. The **G)** CD11b^+^/CCR2^+^ fraction, and **H)** the frequency of CCR2^+^/CD11b^-^ cells are compared among groups (Bonferroni corrected p=0.0125 threshold). For simplicity, in this figure, we referred to CCR2 as -/+ populations, but for the MNPs, it is, in fact, dim/bright populations. Adjusted multiple comparisons two-way ANOVA p values: ¤ <0.05, ¤¤ <0.01, ¤¤¤ <0.001. Healthy control (HC), early PD (EP), late PD (LP) groups.

CD163^+^ infiltrating cells have been observed in PD models and *post-mortem* brains of PwP (Harms et al., 2017; Pey et al., 2014a; Tentillier et al., 2016), and CCR2 has been related to Mo infiltration during α-syn neurodegeneration (Harms et al., 2018). Therefore, we investigated the phagocytic CD163^+^ cells for CCR2 co-expression, regardless of MNP subtyping (**Fig.3D**/**Supp.Fig.3E**). A two-way ANOVA analysis and a post hoc Tukey’s multiple comparisons analysis revealed a significant stage-related change of CD163^+^/CCR2^+^ MNPs, which increased in early PD compared to HCs (**Fig.3E**). We then examined the CD11b^+^/CCR2^+^ MNPs co-expressing both migration markers, of which the majority also are CD163^+^ (**Fig.3F**/**Supp.Fig.3F**). MNPs from early PD had more “triple-positive” cells compared to HCs (and a trend vs. late PD p=0.059) (**Fig.3G**). The early increase remained after covariate adjustment, even though LEDD also had a significant relation to this population. Conversely, both early and late PD showed fewer CCR2^+^ MNPs lacking CD11b (CD11b^-^/CCR2^+^) vs. HCs (**Fig.3H**) that remained significant after covariate adjustment (age and LEDD). Altogether, PwP showed an increased expression of CD163 in MNPs at all stages, and an early expansion of the CD163^+^ cells co-expressing CCR2 and CD11b.

### 4.8 TLR2 expression increases longitudinally and with H&Y (*Cohort#1A and #1B*)

We conducted the statistical analysis of the eight longitudinal samples (*Cohort#1B*) on the six males only, while the female patient and female control were plotted separately. This revealed a progressive (first vs. last visit) MFI increase of the oligomeric α-syn receptor TLR2 on total MNPs and on each Mo subsets (although for iMos first vs. last visit p=0.052) (**Supp.Fig.6A-D**). When examined by repeated measurements, the TLR2 MFI on ncMos was also significantly increased from baseline sampling to 9-12 months later, with the same trending pattern at 1-8 months (**Supp.Fig.6D**). These TLR2 MFI changes were also reflected in the cross-sectional *Cohort#1A* when comparing PwP of H&Y ≤2 vs. >2 (**Fig.4**), showing increased TLR2 MFI on iMos, ncMos, and DCs. Thus, increased TLR2 on MNPs is associated with disease progression and motor worsening. This was not an artifact due to storage time since TLR2 MFI was unrelated to samples’ freezer storage time (not shown).

**Figure 4:**
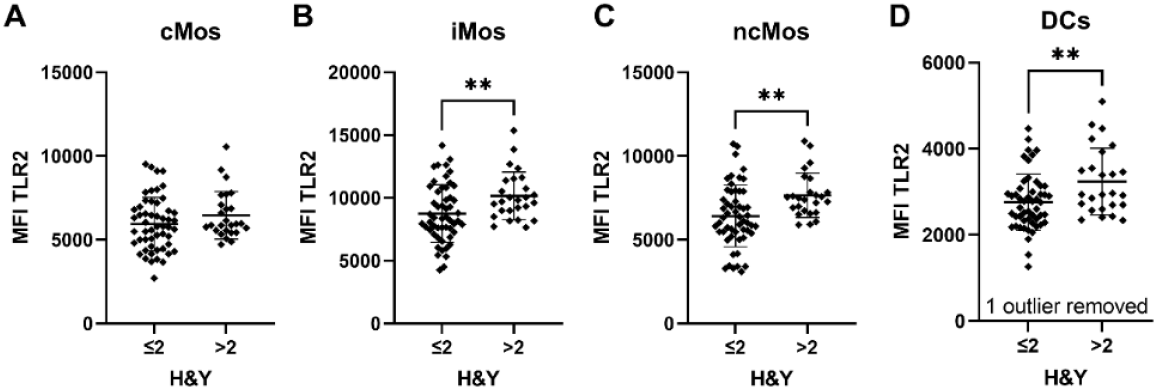
TLR2 expression on mononuclear phagocyte subtypes increases with severity of motor symptoms. Median fluorescents intensity (MFI) of TLR2 on **A)** classical (cMos), **B)** intermediate (iMos), and **C**) non-classical (ncMos) monocytes as well as **D**) dendritic cells (DCs) in patients with Hoehn and Yahr (H&Y) scores ≤2 vs. those with H&Y >2. Non-parametric unpaired t-test, ** p<0.01.

## 5. Discussion

This study examined MNP and NK cell frequencies and their expression of relevant markers in PwP at early- and late-stages since the PD diagnose in a cross-sectional cohort (#1A). We observed that PD is associated with significant alterations in CD163, migration markers (CCR2 and CD11b), and the HLA-DR expressions. Furthermore, PwP showed stage-related alterations in MNP subtypes and receptors levels compared to HCs (summarized in **Fig.5)**. Some changes in the MNPs were sex-specific or were correlated with PD symptoms, supporting an association between peripheral innate immune cells and disease pathology. More specifically, we found that the disease was associated with an increase of iMos in late stages and an early and long-lasting decrease of DCs, with fewer DCs correlated with hyposmia. When comparing PD stages, the pattern suggests an enriched cMos in early vs. late PD, with a mirrored expansion of ncMo in late PD. Notably, these alterations were related to the increased frequency and expression of the CD163 scavenger receptor and the migration markers CCR2 and CD11b, especially in early PD. In contrast, increased HLA-DR expression was more pronounced in late PD. CD16 expression on mature NK cells was also increased in PwP, with a different temporal development according to sex. The only downregulated immune marker in this PD cohort was TLR4, decreasing in females with early PD (**Fig.5**).

**Figure 5:**
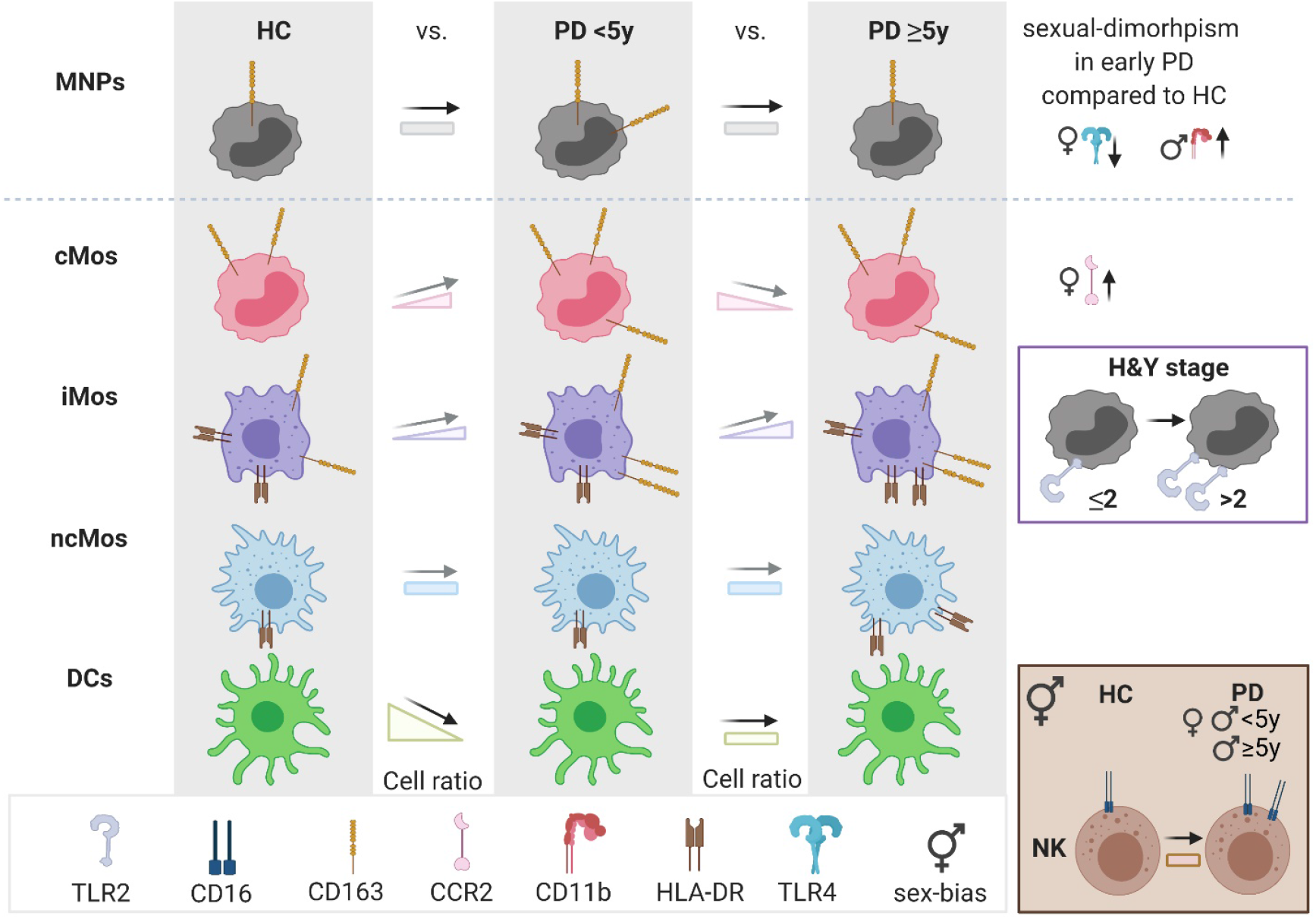
Schematic illustration of the stage-related changes of peripheral blood mononuclear cells in PD. Summary of study findings based on flow cytometry on peripheral mononuclear cells (PBMCs) from people with Parkinson’s Disease (PD) for less than 5 years (<5y), or 5 years and above (≥5y), and healthy control (HC) subjects. Changes in cell type frequencies (in the cell ratio columns: black arrows for significant findings and gray arrows for trends) and surface receptor expressions (drawings on each cell type - for simplicity, CD163 on MNPs refers to percentages of cells positive for this receptors and not the expression) are shown for the innate immune cells in the PBMC pool: for the overall population of TLR2^+^ (inclusion marker) mononuclear phagocytes (MNPs) and uniquely for each subtype of monocytes (classical (cMos), intermediate (iMos), and non-classical (ncMos)), dendritic cells (DCs), and (bottom right frame) the TLR2^-^/CD56^dim^ mature natural killer (NK) cells. NK cells had an increase of CD16 expression in PD with a timing difference for the sexes. Sex differences (far right column) were also observed for the expression levels of TLR4, CCR2, and CD11b with arrows indicating the direction of the change when comparing (early) PD vs. HC. The inclusion marker for MNPs, TLR2 expression, did not change according to disease duration but to Hoehn and Yahr stage (H&Y) in all MNPs except cMos. The illustration was made using BioRender.com.

Interestingly, fewer CD11b^-^/CCR2^dim^ (and ncMos), more CCR2^bright^ MNPs (and cMos) (all in late PD), and increased CCR2 expression (in females), as well as higher MNP viability, were all related to worse cognition in PwP. This suggested that Mo infiltration capacity was associated with cognition; accordingly, when the cohort was divided according to MoCA scores, both the expression level of CCR2 (females only) and CD11b on iMos were higher in patients with cognitive impairment. Furthermore, we showed an upregulation of the α-syn sensor, TLR2, in patients with an H&Y score above 2. This was reflected in increased TLR2 expression in follow-up visits in the longitudinal cases. Altogether, we showed peripheral innate immune changes during early PD with increased cells and proteins related to immune activation, phagocytosis, and extravasation that correlated with symptoms. While some of these changes were maintained when the disease duration extended beyond five years, late PD, many faded maybe as a sign of exhaustion or maybe due to the cells leaving blood to infiltrate tissue. Remarkably, some changes were sex-dependent, suggesting sex-differential immune response during the PD neuroimmune process. Therefore, our study warrants future sex-stratified longitudinal studies.

The distribution of Mo subtypes has previously been investigated with varying results. Some reported no changes (Alvarez-Luquin et al., 2019; da Silva et al., 2016; Drouin-Ouellet et al., 2014; Schlachetzki et al., 2018; Schroder et al., 2018). One of these studies included iMos in the ncMos gate (all CD16^+^), which might explain their increased “ncMos” in CSF from PwP (Schroder et al., 2018). In contrast, another study, including iMos in the cMos gate (all CD14^+^), reported increased cMos and decreased ncMos in PwP vs. controls (Grozdanov et al., 2014). Moreover, a recent study on 41 early PD cases described increased cMos, decreased iMos, and ncMos driven by patients with a high risk of dementia (Wijeyekoon et al., 2020b). Notably, none of these studies analyzed the DCs. The lack of consensus might be related to differences in the stages and types of PD, medication, comorbidities, sex, and flow gating strategy (Marquez et al., 2020; Umehara et al., 2020). As a unique approach, we did not rely only on size or granularity for the MNP selection.

Instead, we pre-gated MNPs using TLR2. The sporadic PD cases included were L-DOPA treated (with few(Yan et al., 2021) exceptions, **Table 1**), sex-balanced, and with a similar prevalence of anti-inflammatory treatment as HCs (**Supp.Table 2**). This approach and the early/late division revealed changes related to PD stages, i.e., disease duration, with a pattern, although not passing the significance threshold, of contraction of the cMos in late vs. early PD. This apparent early cMos expansion is in line with our previous observation of increased cMos in patients with idiopathic REM sleep behavioral disorder (iRBD), considered prodromal PD (Farmen et al., 2021). We observed, however, a late-stage increase of iMos, agreeing with a recent work showing increased iMos in PwP (Thome et al., 2021). Although in their cohort, it was driven mainly by patients at H&Y stage 4, and without information on disease duration, making it difficult to compare with our study directly (Thome et al., 2021). We observed a decrease in DCs, corroborating prior reports of reduced DCs in PwP vs. HCs (Alvarez-Luquin et al., 2019; Ciaramella et al., 2013; Goldeck et al., 2016). We speculate that this reduction reflects a DC migration to the (secondary lymphoid) tissue for Th1/Th17 priming/activation. Accordingly, decreased naïve T cells with increased memory T-cells (Yan et al., 2021) and a Th1/Th17 bias have been reported in PwP (Storelli et al., 2019).

Previously, we observed increased CD56^dim^ mature NK cells in iRBD patients (Farmen et al., 2021), and others reported increased NK cells and NK activity in PwP (Mihara et al., 2008; Niwa et al., 2012). Here, PwP had mature NK cells with increased activation as revealed by higher CD16 expression that occurs early in both sexes but remains late in males PwP. Increased CD16 might suggest an enhanced antibody-dependent cellular cytotoxicity with a differential temporal evolution in each sex. Interestingly, IgG deposition on neurons in close proximity to CD16^+^ (FcγRIII) lymphocytic cells has been shown in *post-mortem* PD brains (Orr et al., 2005). Hence, the increased CD16 expression on NK cells might contribute to adaptive immunity-mediated neurodegeneration. However, future functional analyses are needed to evaluate this hypothesis.

Further suggesting a role for adaptive immunity, we observed increased expression in HLA-DR, which presents antigen-peptides to Th-cells, thus linking innate and adaptive immune components. Increased HLA-DR/MHC-II expression was found on Mos in the CSF from PwP (Fiszer et al., 1994), in *post-mortem* PD brains (Imamura et al., 2003a; McGeer et al., 1988), and PD models (Barkholt et al., 2012; Harms et al., 2013; Thomsen et al., 2021). Correspondingly, we found HLA-DR expression increased on iMos and ncMos in late PD not in early; in agreement with two recent papers showing similar HLA-DR increase (Thome et al., 2021), and one reporting no changes in early PD (Wijeyekoon et al., 2020b). Moreover, we previously observed decreased HLA-DR expression on RBD patients’ MNPs (Farmen et al., 2021), suggesting that HLA-DR expression might be differentially modulated in each disease stage (prodromal, early, and late). The increase in HLA-DR expression might be related to the already mentioned decrease in naive T-cells and expansion of memory CD4 lymphocytes observed in PwP (Yan et al., 2021). A note of caution: this HLA-DR upregulation was also associated with L-DOPA doses, which increase with disease duration and as a consequence of the disease progression in PwP. Thus, it is unclear whether this HLA-DR upregulation is indicative of a PD-related increased antigen presentation capacity or a molecular event induced by the L-DOPA. This will require further investigation.

We and others have shown that MNP-related changes seem particularly relevant for cognition in PD (Ciaramella et al., 2013; Nissen et al., 2021; Wijeyekoon et al., 2020b). Elevated CCR2 expression is a signature of immature Mos marked for migration to blood/tissue. Increased Mo-precursor cells/cMos expressing CCR2 have been described in PD (Funk et al., 2013; Patel et al., 2017), and increased leukocyte CCR2 mRNA expression is associated with cognitive decline (Harries et al., 2012). Here, we showed increased CD11b (in males) and CCR2 expression (in females) and increased CCR2^+^/CD11b^+^ and CCR2^+^/CD163^+^ double-positive MNPs in early PD. Our complex analysis informed about MNPs co-expressing multiple markers contributing to PD rather than CCR2 alone. *Cohort#1A* included mostly PwP with no (MoCA>26) or mild (MoCA=18-26) cognitive impairment, only three had MoCA<18 = (mild) dementia. Despite these relatively high MoCA scores, we observed that the combined absence of both migration markers was associated with better cognition. Accordingly, increased CCR2^+^ MNP percentage (in late PD) or higher CCR2 expression (in females) correlated with impaired cognition. Similarly, we observed that an imbalance of Mo subtypes (increased cMos and reduced ncMos) was associated with worse cognition for the late patients. Furthermore, the iMos’ MFI of CD11b and CCR2 (females only) was higher in those patients with a MoCA score indicative of cognitive impairment (MoCA ≤26). This corroborates that Mos with migratory capacity are associated with cognitive decline in PwP (Nissen et al., 2021; Umehara et al., 2020; Wijeyekoon et al., 2020b). Therefore, indicating that an apparent long-lasting Mo-deviation from healthy levels is disadvantageous for the patients. This deserves further investigation in a cohort with severe cognitive impairment.

Immunological sexual-dimorphism, including sex-divergent Mo aging (Marquez et al., 2020), has been described (Bouman et al., 2004; Sue, 2017). In our study, some sex differences seemed constitutive, such as the higher female TLR4 expression observed in HCs. In contrast, sexual-dimorphism for CCR2 and CD11b was apparent only in PwP. The higher TLR4 expression in female HCs might be particularly relevant due to TLR4’s protective role by clearing α-syn (Choi et al., 2020; Earls and Lee, 2020), which could contribute to the lower PD risk for females. However, prolonged misfolded α-syn-sensing by TLR4 also contributes to (chronic) inflammation and thus to neurodegeneration (Campolo et al., 2019), which might be differently regulated in females and males. The relevance of TLR4 is reinforced by our previous data showing Mo TLR4 expression positively correlating with midbrain immune activation and decreased dopaminergic putaminal transmission in iRBD (male) patients (Farmen et al., 2021). In that iRBD cohort, TLR2 expression was correlated with hyposmia. Moreover, here in our cross-sectional *Cohort#1A*, we detected a significant increased TLR2 expression on iMos, ncMos, and DCs in patients with H&Y scores above two vs. those with lower scores. Likewise, we observed increasing TLR2 expression on all MNPs in our longitudinal case reports (*Cohort#1B*). Although previously reported data on TLRs in other PD cohorts are conflicting (da Silva et al., 2016; Drouin-Ouellet et al., 2014; Wijeyekoon et al., 2020b), our prior data in iRBD (Farmen et al., 2021) and the observations here in PwP confirm the relevant role of TLR2/4 in PD-pathology suggested previously by human, animal, and cellular studies (Harms et al., 2021).

TLR activation and other inflammatory events induce shedding of the MNP-specific membrane receptor CD163, forming sCD163 (Moller, 2012). Plasma sCD163 correlates negatively with CD163 MFI, but not with CD163^+^ cell counts (Davis and Zarev, 2005); thus, increased sCD163 paralleled a decreased CD163 MFI in multiple sclerosis (Gjelstrup et al., 2018). With our previous finding of increased serum sCD163 levels in females with PD from another cohort (Nissen et al., 2021), we anticipated decreased CD163 MFI on MNPs from females with PD in *Cohort#1A* (same biobank, different donors). However, we observed significantly increased CD163 MFI in PwP for both sexes, indicating a PD-associated upregulation of CD163 that would parallel the increased shedding in serum (females) and CSF (both sexes) that we described before (Nissen et al., 2021). Although CD163 upregulation was not observed in a cohort of iRBD (male) patients, increased CD163^+^ cell frequency was associated with reduced immune activation in the midbrain and preserved dopaminergic neurotransmission in the putamen, both shown by PET imaging (Farmen et al., 2021). This was suggestive of a protective role of the CD163 population at this prodromal PD stage. Indeed, CD163 expression has previously been associated with alternative activation of MNPs considered of anti-inflammatory character (Fischer-Riepe et al., 2020).

The PD-related CD163 upregulation and co-expression with CD11b and CCR2 suggest CD163^+^ cell infiltration into CNS. Infiltrated CD163^+^ cells are found in the *post-mortem* brains of PD models and PwP (Harms et al., 2017; Pey et al., 2014b; Tentillier et al., 2016). The brain infiltration and subsequent activation of the CD163^+^ cells may lead to sCD163 production, in agreement with our previously reported increased sCD163 in CSF from PwP (Nissen et al., 2021). Furthermore, the CSF-sCD163 levels in PwP were correlated with markers of angiogenesis and chemotaxis, such as ICAM-1 (CD11b agonist) and CCL2 (CCR2 ligand)), CSF-α-syn, Tau, and cognition (Nissen et al., 2021). Altogether, our current and prior data suggest that CD163^+^/CCR2^+^/CD11b^+^ cells seem to infiltrate the brain of PwP, differentiate into macrophages, and influence the CNS inflammatory status and neurodegeneration. However, the CD163 receptor’s specific role in PD remains to be determined.

The fewer HCs might limit the present study. Additionally, alternative separation based on PD subtypes or progression rates (Wijeyekoon et al., 2020b) might challenge our early/late-division based on disease duration. Nevertheless, this division revealed otherwise hidden differences, which could explain previous inter-study MNP inconsistencies in PD. Moreover, this did not preclude us from associating changes with scores related to disease severity (MoCA and H&Y).

## 6. Conclusion

Altogether, our study suggests that the PD immune response differs with disease duration/stages by activation in early PD and some degree of immune exhaustion or stabilization emerging at late-stage. We show a longitudinal increase of TLR2 on MNPs in PwP, related to motor worsening by H&Y in the cross-sectional cohort. Our data indicate stage (disease duration)-associated alterations in the innate immune compartment, particularly in the phagocytic CD163^+^ migration competent CCR2^+^/CD11b^+^ cMos and iMos, but for all TLR2^+^ MNPs in general. We observed increases of innate immune cells and receptors, especially in early PD, while HLA-DR was upregulated mainly in late PD. We also showed significantly increased CD16 expression on mature NK cells, relevant for antibody-dependent cytotoxicity, with a different temporal evolution in males and females. Further highlighting the sex dimorphism observed in PD, we also observed sex-related TLR4, CCR2, and CD11b differences. Interestingly, many of these changes were associated with PD symptoms: such as that of TLR2 with the motor symptoms, DCs with hyposmia, plus CCR2 and CD11b with cognition. Altogether, our findings show alterations in the peripheral immune system in PD related to the disease duration and sex and highlight a need to study CD163^+^ MNPs further, focusing on the role of cMos and iMos in PD.

## Data Availability

The data supporting the conclusions are included within the article and its additional file.

## 7. Declarations

### 7.1 Ethics approval and consent to participate

The study was approved by the Ethics Committee of the University of Tuebingen (Germany) committee (480/2015BO2). All participants providing written informed consent.

### 7.2 Availability of data and materials

The data supporting the conclusions are included within the article and its additional file.

### 7.3 Financial disclosure during the past 12 months

K. Brockmann has received a research grant from the University of Tuebingen (Clinician Scientist), the dPV, the MJFF, and the German Centre for Neurodegenerative Diseases (DZNE, MIGAP); travel grants from the Movement Disorders Society, and speaker honoraria from Abbvie, Lundbeck, UCB, and Zambon.

M. Romero-Ramos serves on the editorial board of Brain Research and npj Parkinson’ disease, and receives research support from Novo Nordisk Foundation, Aarhus University Research Foundation, the Danish Medical Research Council, the Danish Parkinson Foundation, Desiree and Niels Ydes Foundation, and the Michael J Fox Foundation.

SKN, KF, MC, CS, and DG report no disclosures.

### 7.4 Authors’ contribution

SKN: Study design, flow of *Cohort#1A*, data analysis, wrote the first draft; KF: Flow and analysis of *Cohort#1B*; MC: Flow; CS: Coordinated sample selection and collected biobank archive info; DG: isolated PBMCs; KB: Supervised the biobank; MRR: Developed the concept and designed the study. MRR & SKN: Interpreted the data and wrote jointly the final version. All authors critically revised the manuscript.

## 7.5 Acknowledgments

We acknowledge the invaluable technical help provided by Gitte Ulbjerg Toft (Department of Biomedicine, Aarhus University). Samples were obtained from the Neuro-Biobank of the University of Tuebingen, Germany (https://www.hih-tuebingen.de/en/about-us/core-facilities/biobank/), which is supported by the local University, the Hertie Institute and the DZNE. Flow cytometry was performed at the FACS Core Facility, Aarhus University, Denmark.

