## supplementary information for "Changes in CD163+, CD11b+, and CCR2+ peripheral monocytes relate to Parkinson’s disease and cognition"

### Supplementary material

**Sample collection**

Based on the local biobank standardized protocol, one to two 9 ml vials of EDTA blood were collected with similar collection time points. Peripheral blood mononuclear cells (PBMCs) were isolated using Ficoll-Hypaque gradient on the collection day. $10^6$-$10^7$ PBMCs/ml in RPMI-1640 medium with 10% DMSO (Serva) and 20% FBS (Sigma) were frozen at -80°C, and subsequently stored at -135°C or colder. Samples were shipped at dry ice when transferring between Germany and Denmark, where they were stored at -135°C until processing.

**Flow cytometry on *Cohort#1(A&B)* and DC identification**

PBMCs were thawed in 10 mL preheated RPMI 1640 Glutamax (Gibco) with 1% penicillin/streptomycin, washed, and counted. One million cells were lived/dead stained using 0.1 µL LIVE/DEAD™ Fixable Near-IR Dead Cell Stain Kit, for 633 or 635 nm excitation (Invitrogen, Thermo Fisher Scientific) in 80 µL DPBS (Biowest), followed by a DPBS wash, and subsequent blocking in 100 µL 10% heat-inactivated (HI) human AB Serum (H4522 Sigma) in DPBS. Cells were stained with anti-human cell surface fluorochrome-conjugated antibodies according to **Supp.Table 3** in a total volume of 100 µL DPBS with 1% Albumin fraction V, from bovine serum (Merck). After additional washing and fixation in 0.9% formaldehyde (SigmaAldrich) in DPBS, the cells were stored at 4°C for 1-3 days and run on an LSR Fortessa flow cytometer (BD) equipped with a 405 nm, 488 nm, 561 nm, and 640 nm laser. A compensation matrix was calculated using single stains on OneComp eBeads (Thermo Fischer Scientific) and ArC Armine reactive compensation bead kit (Invitrogen) for antibodies and live/dead stain, respectively. Daily cytometer voltage settings were confirmed using SPHERO™ Rainbow calibration Particles (BD Bioscience), reassuring the same MFI in all open channels for at least three peaks. Beforehand, all antibodies were titrated on at least five different dilutions of single stains. Proper compensation and gating were confirmed on fluorescence minus one (FMO) samples. Patient subgroups and controls were evenly distributed in different experiments. For each experiment, aliquots from the same control donor were stained to test any day-to-day variation and antibody lot number variation. Antibody lot differences affected TLR4; thus, the

analyses only include the 66 samples where the same TLR4 antibody lot was used. FSC files were analyzed blinded using FlowJo V10 with the following gating strategy (**Supp.Fig.3**): Single cells were identified on an FSC-A vs. FSC-H followed by SSC-A vs. SSC-H, debris was excluded on a FSC-A vs. SSC-A plot. Cytometer stability was gated on time vs. CD163 PE-A, with subsequent exclusion of dead cells on live/dead vs. SSC-A. Monocytes/dendritic cells (DCs), thus mononuclear phagocytes (MNPs) ($TLR2^{+}$), the precursor NK cells ($CD56^{bright}/TLR2^{-}/CD16^{-}$), and mature NK cells ($CD56^{dim}/TLR2^{-}/CD16^{+/-}$) were identified on CD56 ECD-A vs. TLR2 APC-A with CD16 and CD11b heat-mapping as guidance. From the MNP gate, several different populations were identified: CD14 BV421-A vs. CD16 PC7-A with HLA-DR heat-mapping (based on its high monocytic discrimination index (1, 2)) was used to identify MNP subtypes (classical- (cMos), intermediate- (iMos), and non-classical monocytes (ncMos) as well as DCs); CD163 PE-A vs. CCR2 FITC-A (fixed gating); and CD11b SB780-A vs. CCR2 FITC-A with CD163 heat-mapping (for the CCR2/CD11b $double^{+}$ population, the majority was also $CD163^{+}$). Frequencies of single maker expression on MNPs and other subpopulations were gated (fixed) against SSC-A. Median fluorescence intensity (MFI) was calculated on gates with only a single peak/homogeneous populations, or on priory positive/bright-gated cells as indicated.
A t-distributed stochastic neighbor embedding (t-SNE) plot was conducted on ten concatenated samples of live cells from all patient and control groups to confirm gating strategy and explore co-expression of different receptors.

To examine the DC distribution, we used another antibody panel (**Supp.Table 3**) and gating strategy (**Suppl.Fig4**) on three control samples obtained from blood donations buffy coats at the department of Clinical Biochemistry at Aarhus University Hospital, Skejby, Denmark. A similar gating approach as described above was used to reach MNP subtyping (Q1-4), except no gate on time. From the live cell gate, conventional DC type 1 (cDC1) and plasmacytoid (p)DCs were identified on dot plot with TLR2 APC-A vs. CD141 BV510-A or vs. CD303 PerCP-eFlour710-A, respectively (2). With a fraction of B-cells also being CD1c positive, the cDC type 2 (cDC2) was identified from the MNP gate on a CD1c PE-A vs. SSC dot plot. cDC2 was back-gated on MNP subtypes (Q1-4), and to the MNP main gate together with cDC1 and pDC back-gating. From the Q4 gate on the CD14 BV421-A vs. CD16 PC7-A MNP subtyping dot plot, CD1c PE-A vs. CD11c FITC-A was used to define the actual DCs within this gate.

**Supplementary Table 1: Clinical details for the longitudinal *Cohort#1B***

| Code+visit | Month from V1 | Age at onset | Age at visit | Disease duration | LEDD | H&Y | UPDRS III | MoCA |
|---|---|---|---|---|---|---|---|---|
| **EPM21.v1** | - | 66-70 | 66-70 | 2 | 100 | 1 | 7 | 29 |
| **EPM21.v2** | 6 | - | - | 3 | 100 | 1 | 10 | 29 |
| **EPM21.v3** | 12 | - | - | 3 | - | 1 | 10 | 28 |
| **EPM23.v1** | - | 71-75 | 76-80 | 4 | - | 2 | 30 | 24 |
| **EPM23.v2(L)** | 11 | - | - | 5 | 643 | 2 | 41 | 27 |
| **LPM21.v1** | - | 60-65 | 70-75 | 7 | 965 | 3 | 21 | 29 |
| **LPM21.v2** | 6 | - | - | 8 | 1213 | 3 | 28 | 25 |
| **LPM21.v3** | 12 | - | - | 8 | 1213 | 2 | 11 | 26 |
| **LPM21.v4** | 16 | - | - | 8 | 1307 | 2 | 23 | 28 |
| **LPM22.v1** | - | 56-60 | 66-70 | 9 | 920 | 2 | 37 | 28 |
| **LPM22.v2** | 6 | - | - | 10 | 900 | 3 | 25 | 28 |
| **LPM22.v3** | 12 | - | - | 10 | 900 | 2 | 19 | 30 |
| **LPM23.v1** | - | 46-50 | 61-65 | 15 | 1454 | 2 | 29 | 28 |
| **LPM23.v2** | 1 | - | - | 15 | 1087 | 2 | 42 | 30 |
| **LPM24.v1** | - | 51-55 | 56-60 | 5 | 420 | 3 | 70 | 26 |
| **LPM24.v2** | 5 | - | - | 6 | 500 | 2 | 59 | 28 |
| **LPM24.v3** | 11 | - | - | 6 | 780 | 3 | 63 | 26 |
| **LPF21.v1** | - | 56-60 | 66-70 | 9 | 665 | 3 | 25 | 27 |
| **LPF21.v2** | 6 | - | - | 10 | 715 | 3 | 30 | 28 |
| **HCF15.v1** | - | - | 66-70 | - | - | - | - | 28 |
| **HCF15.v2** | 6 | - | - | - | - | - | - | 26 |

Patients with early- (<5 years since diagnosis) or late-stage (≥5 years since diagnosis) sporadic Parkinson’s disease (PD) and a healthy control (HC) of who the biobank provide samples from multiple visits: Early PD male (EPM) (with progression to late (L) disease status), late PD male (LPM), late PD female (LPF), and HC female (HCF). Time at sampling from baseline/visit (v)1 is shown in months. Due to few individuals with longitudinal samples available, five years age ranges are shown for age-at-onset and age-at-visit/sampling time. Disease duration is shown in years. Clinical information, if available at sampling time, is indicated for L-dopa equivalent daily dose (LEDD), Unified Parkinson's Disease Rating Scale three (UPDRS III), Hoehn and Yahr (H&Y score), and the Montreal Cognitive Assessment (MoCA) score. Sample coding was randomly assigned in the laboratory. Neither doctors nor patients are familiarized with their code number.

#### Supplementary Table 2: Non-dopaminergic treatment related to inflammation and cognition for (*Cohort#1A*)

| | HC | PD | P |
|---|---|---|---|
| **Individuals receiving anti-inflammatory** (AI) **treatment without diagnosis: total** | **2** | **9** | ns |
| - Antihistamine | 1 | 0 | |
| - NSAID (Ibuprofen) | 1 | 4 | |
| - NSAID (Diclofenac) | 0 | 2 | |
| - Corticosteroid (Astonin) | 0 | 1 | |
| - Anti-microbe (Aciclovir, Quensyl) | 0 | 2 | |
| **Total anti-inflammatory treatment** | **2** | **9** | ns |
| **Diabetic treatment** (DT) (Glimepirid, Metformin, Siofor, Novorapid, rotaphane) | 0 | 3 (only) 3 (+ND) 1 (+AI) **7 total** | ns |
| **AI + DT** | **2** | **16** | ns |
| **Neurological drugs other than dopaminergic treatment** (ND) (Amitriptylin, Cipramil, Citalopram, Clozapin, Cymbalta, Elontril, Exelon, Gabapentin, Laif, Lithium, Mirtazapin, Orfiril, Quetiapin, Remergil, Rivastigmin, Rivotril, Seroquel, Sertralin, Stangyl, Trimipramin, Valproat, Venlafaxin) | 0 | 21 (only) 3 (+DI) 5 (+AI) **29 total** | **0.0001** |
| **Hormone related treatment** | **1** | **1** | ns |
| - Evista (selective estrogen receptor modulator) | 1 | 0 | |
| - Estragest | 0 | 1 | |
| **Treated for any of the above** (overlap)**:** | **5** (2) | **48** (9) | **0.0001** |
| **No relevant treatment or diagnosis** | **24**/29 | **30**/80 | **0.0001** |

The tow-tailed P values are shown for Fisher’s exact test. Nonsteroidal anti-inflammatory drug (NSAID).

#### Supplementary Table 3: Fluorochromes and antibodies used for flow staining

| Panel | Antigen | Fluorophore | Isotype | Clone | Host | µL/ 100 | Company |
|---|---|---|---|---|---|---|---|
| Cohort#1 | CD163 | PE | IgG1 | MAC2-158 | Mouse | 1 | IQProducts |
| Cohort#1 | TLR4 | PerCP | IgG2a | 610015 | Mouse | 10 | Novus, R&D systems |
| Cohort#1 | CD192 (CCR2) | FITC | IgG2a κ | K036C2 | Mouse | 5 | Biolegend |
| Cohort#1 | CD11b | Super Bright 780 | IgG1 | ICRF44 | Mouse | 1.25 | eBioscienceTM Thermo Fischer Scientific |
| Cohort#1 / DC | CD14 | BV421 | IgG2b | Mϕp9 | Mouse | 3 | BD Horizon |
| Cohort#1 /DC | CD16 | PC7 | IgG1 | 3G8 | Mouse | 2.5 | Beckman Coulter |
| Cohort#1 /DC | TLR2 (CD282) | APC | recombinant | REA109 | | 2 | MACS Miltenyi Biotec |
| Cohort#1 /DC | HLA-DR | BV650 | IgG2a κ | L243 | Mouse | 2.5 | Biolegend |
| Cohort#1 /DC | CD56 | ECD | IgG1 | N901 | Mouse | 2 | Beckman Coulter |
| Cohort#1 /DC | LIVE/DEAD™ Fixable Near-IR Dead Cell Stain Kit | 633/ 635nm | | | | 0.1 | Invitrogen, Thermo Fisher Scientific |
| DC-panel | CD141 | BV510 | BALB/c IgG1,K | 1A4 | Mouse | 5 | BD Horizon TM, BD |
| DC-panel | CD303a (BDCA-2) | PerCP-eFlour 710 | IgG2a κ | 201A | Mouse | 5 | Invitrogen |
| DC-panel | CD1c | PE | IgG κ | L161 | Mouse | 5 | eBioscience TM Invitrogen |
| DC-panel | CD11c | FITC | IgG1 κ | B-ly6 | Mouse | 2 | BD Pharming TM |

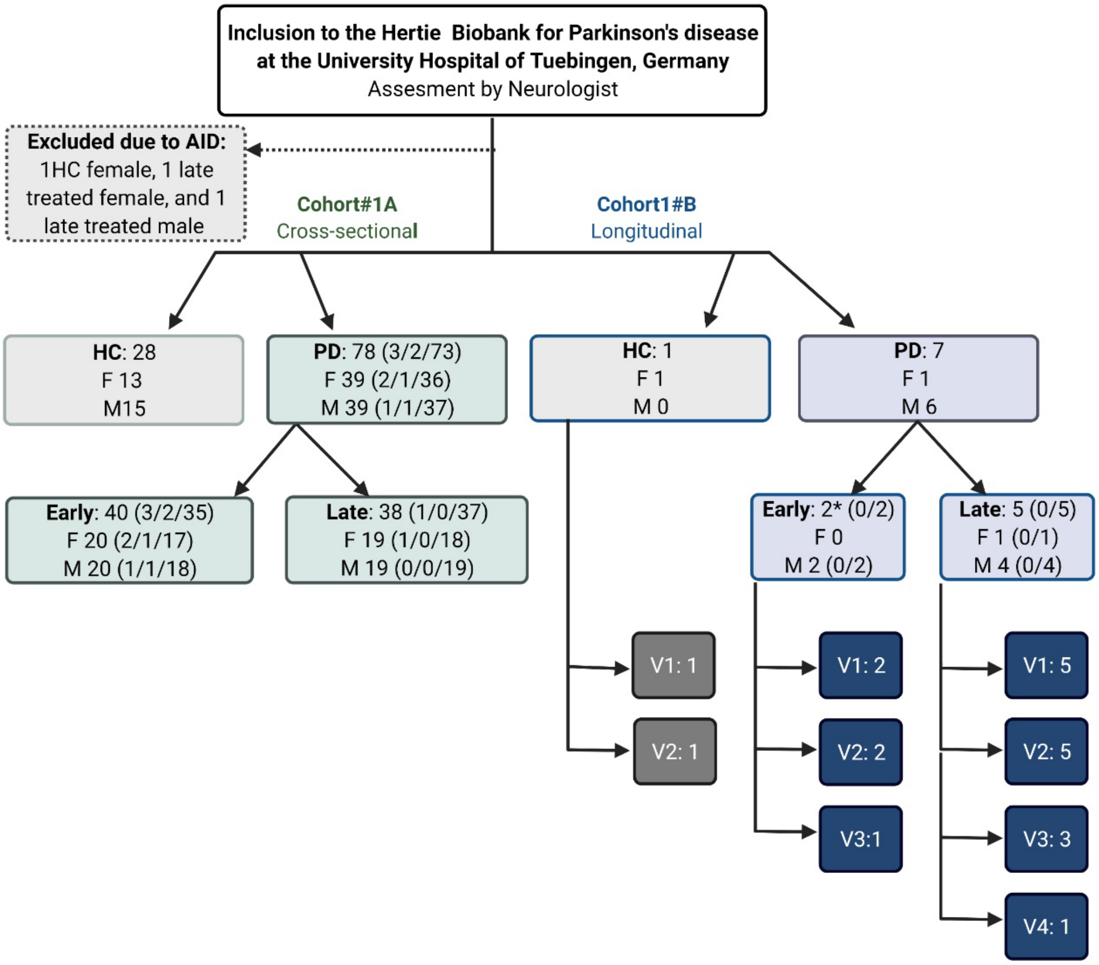


**Supplementary Figure 1: Overview of study participants**

The study includes samples from healthy control (HC) individuals and people with sporadic Parkinson's disease (PD) from the Hertie Biobank for Parkinson's disease at the University Hospital of Tuebingen, Germany. Subjects with autoimmune diseases (AID) were excluded. The cohort was subdivided into a cross-sectional *Cohort#1A* with 28 HC individuals and 78 people with PD, and *Cohort#1B* for case studies of longitudinal samples with various visit (v) numbers. The distribution of females (F) and males (M) are shown with dopaminergic treatment status in brackets: (unknown/untreated/treated).

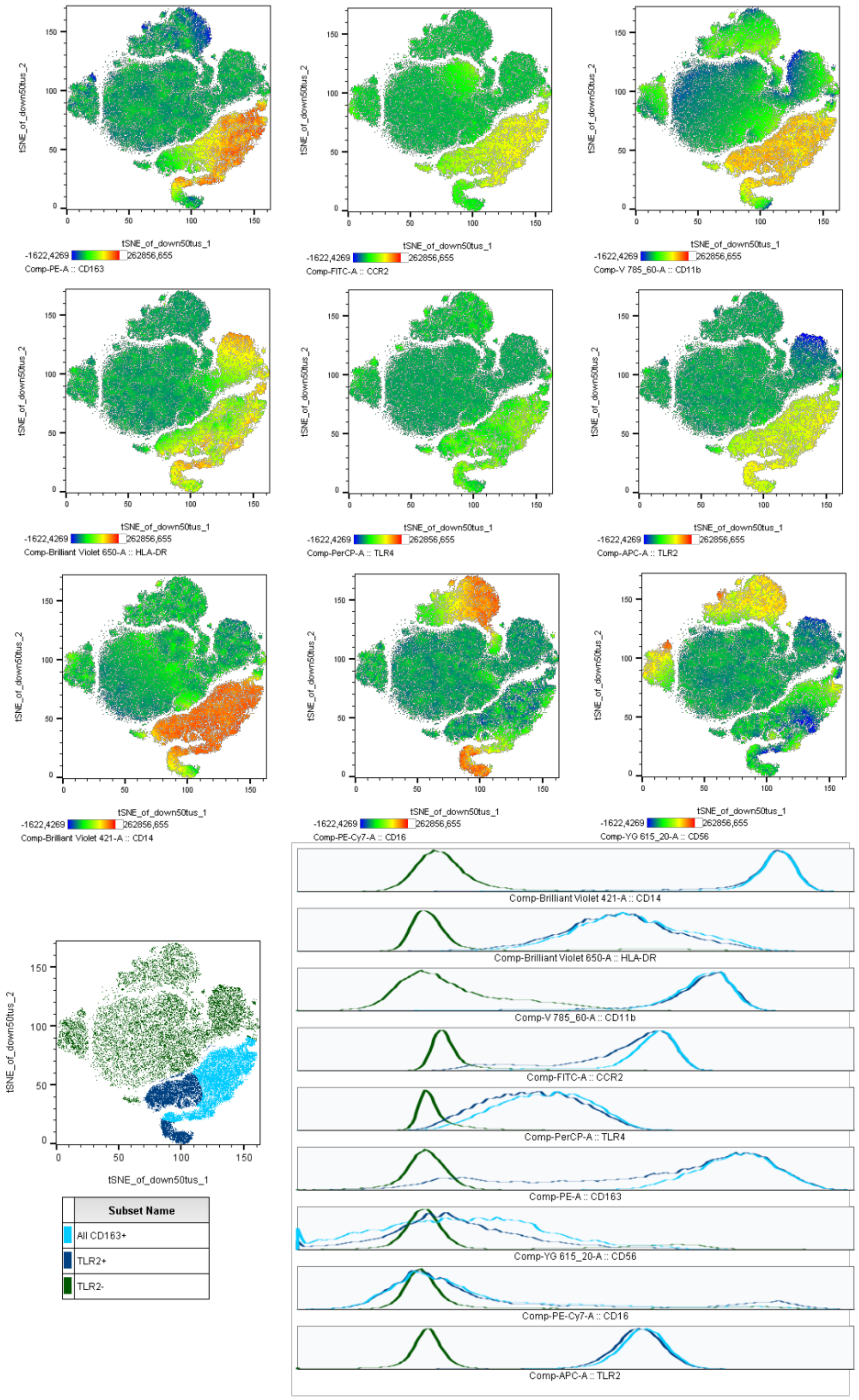


**Supplementary Figure 2. Unraveling TLR2 as a mononuclear phagocyte marker**
t-distributed stochastic neighbor embedding (tSNE) dimensionality reduction plots of ten concatenated samples of live cells from all PD and control groups with expression heat maps of all surface markers; and a dot plot with overlay of all TLR2$^{+}$ cells (dark blue), all CD163$^{+}$ cells (light blue), and TLR2$^{-}$ cells (green); accompanied by histograms of surface makers' expression.

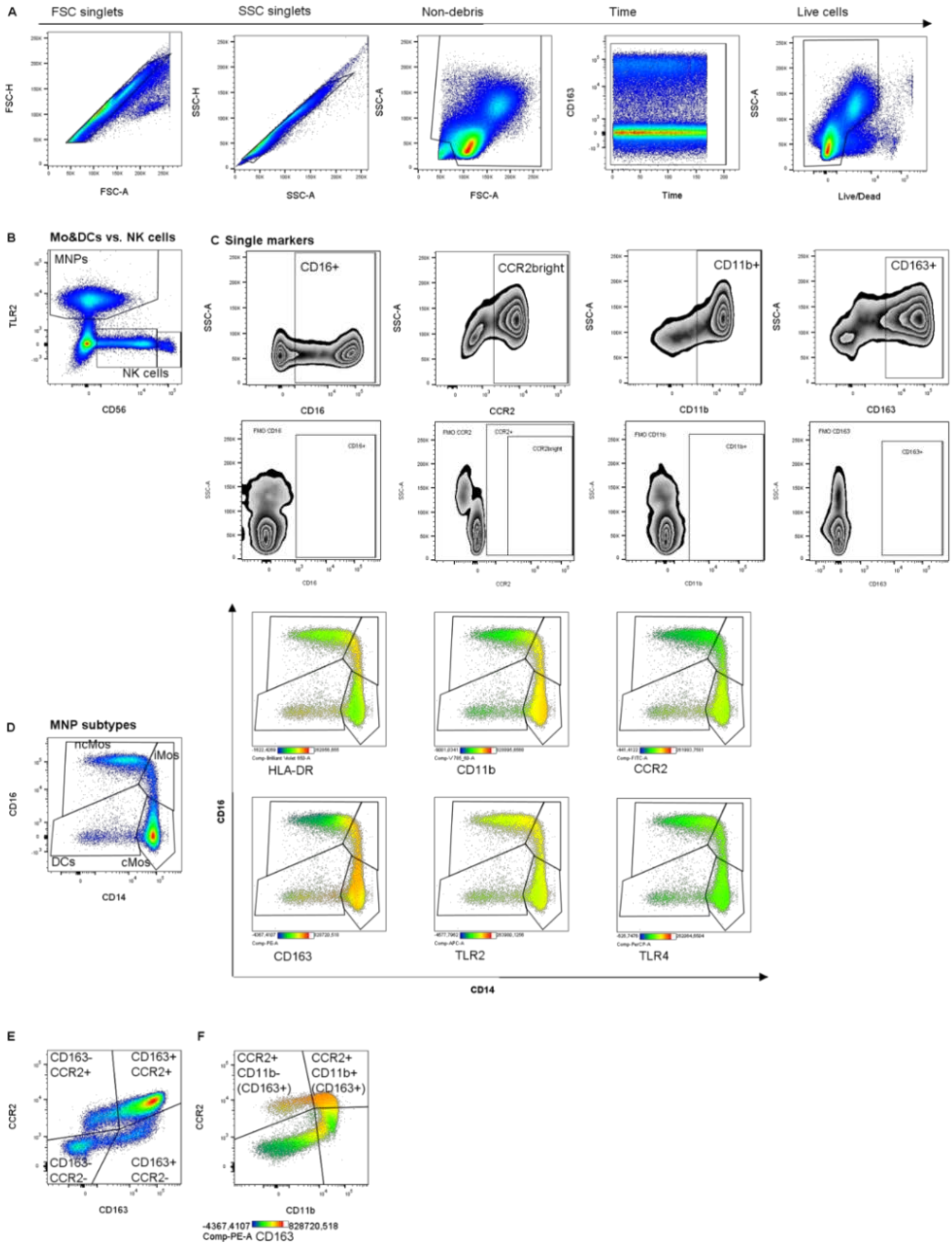
A
FSC singlets
SSC singlets
Non-debris
Time
Live cells
FSC-H
FSC-A
SSC-H
SSC-A
SSC-A
FSC-A
CD163
Time
SSC-A
Live/Dead
B
Mo&DCs vs. NK cells
MNPs
NK cells
TLR2
CD56
C Single markers
CD16+
CCR2bright
CD11b+
CD163+
CD16
CCR2
CD11b
CD163
D
MNP subtypes
ncMos
iMos
DCs
cMos
CD16
CD14
HLA-DR
CD11b
CCR2
CD163
TLR2
TLR4
CD16
CD14
E
CD163-
CCR2+
CD163+
CCR2+
CD163-
CCR2-
CD163+
CCR2-
CCR2
CD163
F
CCR2+
CD11b-
(CD163+)
CCR2+
CD11b+
(CD163+)
CCR2
CD11b
-4367,4107
828720,518
Comp-PE-A CD163

**Supplementary Figure 3: Gating strategy for *Cohort#1A&B***

Identification of **A)** single live cells in the PBMC population based on forward scatter (FSC) and side scatter (SSC) height (H) vs. area (A), exclusion of debris, cytometer performance validation over time, followed by dead cell exclusion. **B)** Mononuclear phagocytes (MNPs), including monocytes (Mos) and dendritic cells (DCs), were identified from the live cell populations as TLR2$^{+}$; and mature and precursor natural killer (NK) cells were gated as TLR2$^{-}$ /CD56$^{dim}$ and TLR2$^{-}$/CD56$^{bright}$, respectively. **C)** Frequencies of single markers were identified using fixed gates against SSC-A for CD16$^{+}$ (mature NK cells only), CCR2$^{bright}$, CD11b$^{+}$, and CD163$^{+}$ when appropriate (upper panel: full stained; lower panel: fluorescence minus one (FMO) controls). **D)** From the MNP gate, classical (cMos), intermediate (iMos), non-classical monocytes (ncMos), and DCs were identified based on CD14 and CD16 expression with gates adjusted based on HLA-DR heat-mapping. Heat maps are shown for other markers to visualize the MFI on the different subtypes. Observed subtype characteristics, which were used to guide further analysis: cMos CD14$^{++}$/CD16$^{-}$/HLA-DR$^{dim}$ (CD11b$^{+}$/CCR2$^{bright}$/CD163$^{bright}$/TLR2$^{bright}$), iMos CD14$^{++}$/CD16$^{+}$/HLA-DR$^{bright}$ (/CD11b$^{+}$/CCR2$^{bright}$/CD163$^{bright}$/TLR2$^{bright}$), ncMos CD14$^{dim}$/CD16$^{++}$/HLA-DR$^{dim}$ (/CD11b$^{-/dim}$/CCR2$^{+}$/CD163$^{dim/-}$/TLR2$^{bright}$), and DCs CD14$^{dim/-}$/CD16$^{-}$/HLA-DR$^{bright}$ (/CD11b$^{-}$/CCR2$^{+}$/CD163$^{Mixed}$/TLR2$^{dim}$). TLR4 was similar in all subtypes; thus, only investigated in the full MNP population. **E)** To investigate the relation of CD163 expression with infiltration/transmigration markers, from the MNP gate in (B), CD163 vs. CCR2 was gated with fixed gates, and **F)** CD11b vs. CCR2 was gated with adjustments made based on CD163 heat-mapping. Median fluorescence intensity (MFI) was measured on all MNP if expressed homogeneously (e.g., TLR4 in all TLR2$^{+}$), or on single populations with high expression of the marker (CCR2$^{bright}$ populations: cMos, and iMos); or into the positive gate only (e.g. of all CD16$^{+}$ gated population). Thus, MFI was not analyzed on all data subsets.

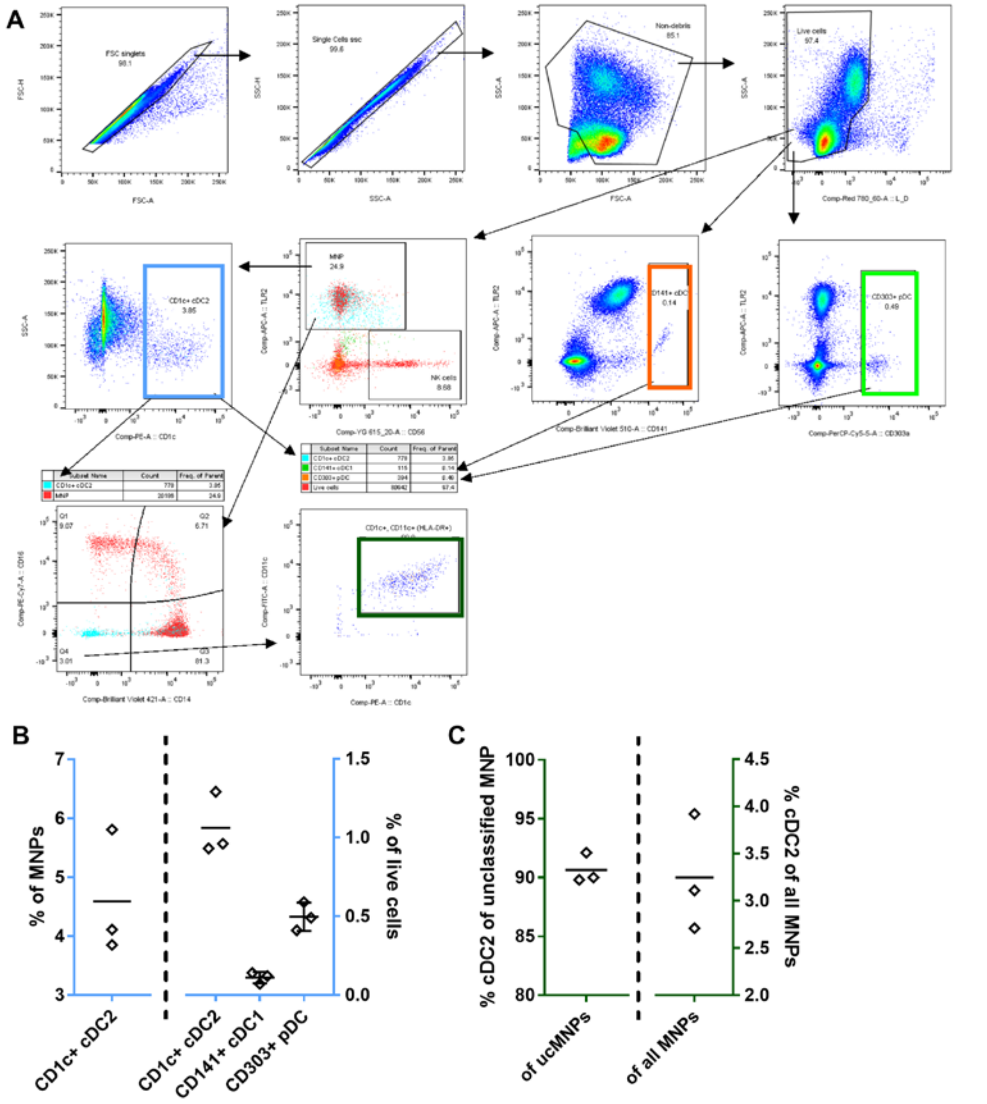


**Supplementary Figure 4: Identification of dendritic cell subtypes**

A control staining for dendritic cells (DCs) was performed on three control donors. **A)** Similar gating approach as described in **Supp.Fig2** was used to reach mononuclear phagocyte (MNP) subtyping (Q1-4) on CD14 vs. CD16; except for no gate on time. From the live cell gate, conventional (c)DC type 1 (cDC1) and plasmacytoid (p)DCs were identified as TLR2$^-$/CD141$^+$ and TLR2$^-$/CD303$^+$, respectively. With a fraction of B-cells (TLR2$^-$) also being CD1c$^+$, the cDC type 2 (cDC2) was identified from the MNP gate on a CD1c vs. SSC dot plot. cDC2 was back gated onto MNP subtypes (Q1-4) and to the MNP main gate together with cDC1 and pDC back-gating. From the Q4 gate on the CD14 vs. CD16 MNP subtyping dot plot, CD1c vs. CD11c was used to define the actual cDC2s within this gate. **B)** DC subtypes as percentages of MNPs or all live cells. **C)** CD1c$^+$/CD11c$^+$ cDC2 percentage of the Q4 gate (termed DCs in *Cohort#1*) and of all MNPs.

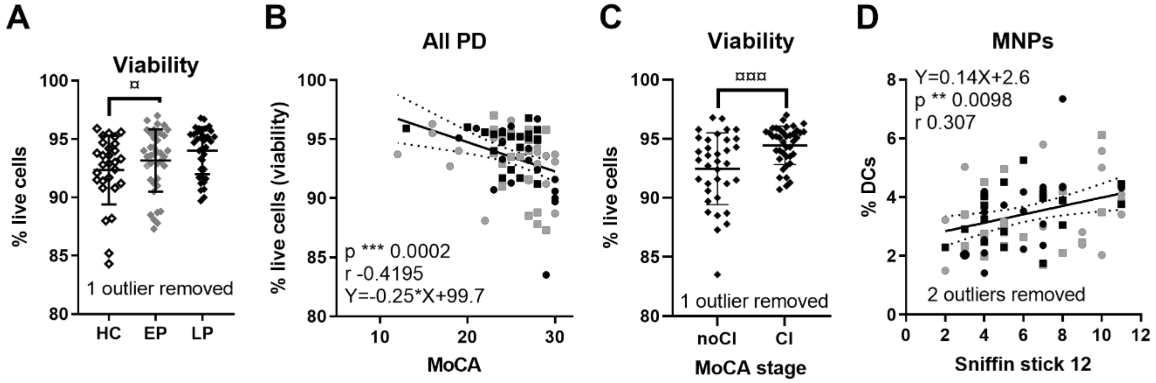


**Supplementary Figure 5: Fewer dendritic cells (DCs) weakly associates with hyposmia (*Cohort #1A*)**

**A)** Frequency of live cells measured in the healthy control (HC), early PD (EP, <5 years since diagnosis), or late PD (LP, ≥5 years since diagnosis) groups, respectively (significance lost after adjustment for age (and LEDD)). One outlier removed from the LP group (ROUT, Q = 0.1%) and compared with two-way ANOVA with Tukey's multiple comparinsons. **B)** Variation in cell viability from PwP (symbols: females round, males square, early gray, late black) correlated negatively with the Montreal Cognitive Assessment (MoCA) scores; Spearman p and r values, and linear regression equation are shown. **C)** Patients were subdivided according to the MoCA stage with score >26 = no cognitive impairment (noCI) and ≤26 = cognitive impairment (CI). One outlier was removed from the CI group. **D)** Pearson correlation and linear regression of the olfaction scores Sniffin' Stick 12 versus the percentage of $CD14^{-/low}/CD16^{-}$ DCs (type cDC2) within the mononuclear phagocytes (MNPs). Two outliers were removed. The linear relation was not affected by age at visit, nor by LEDD.

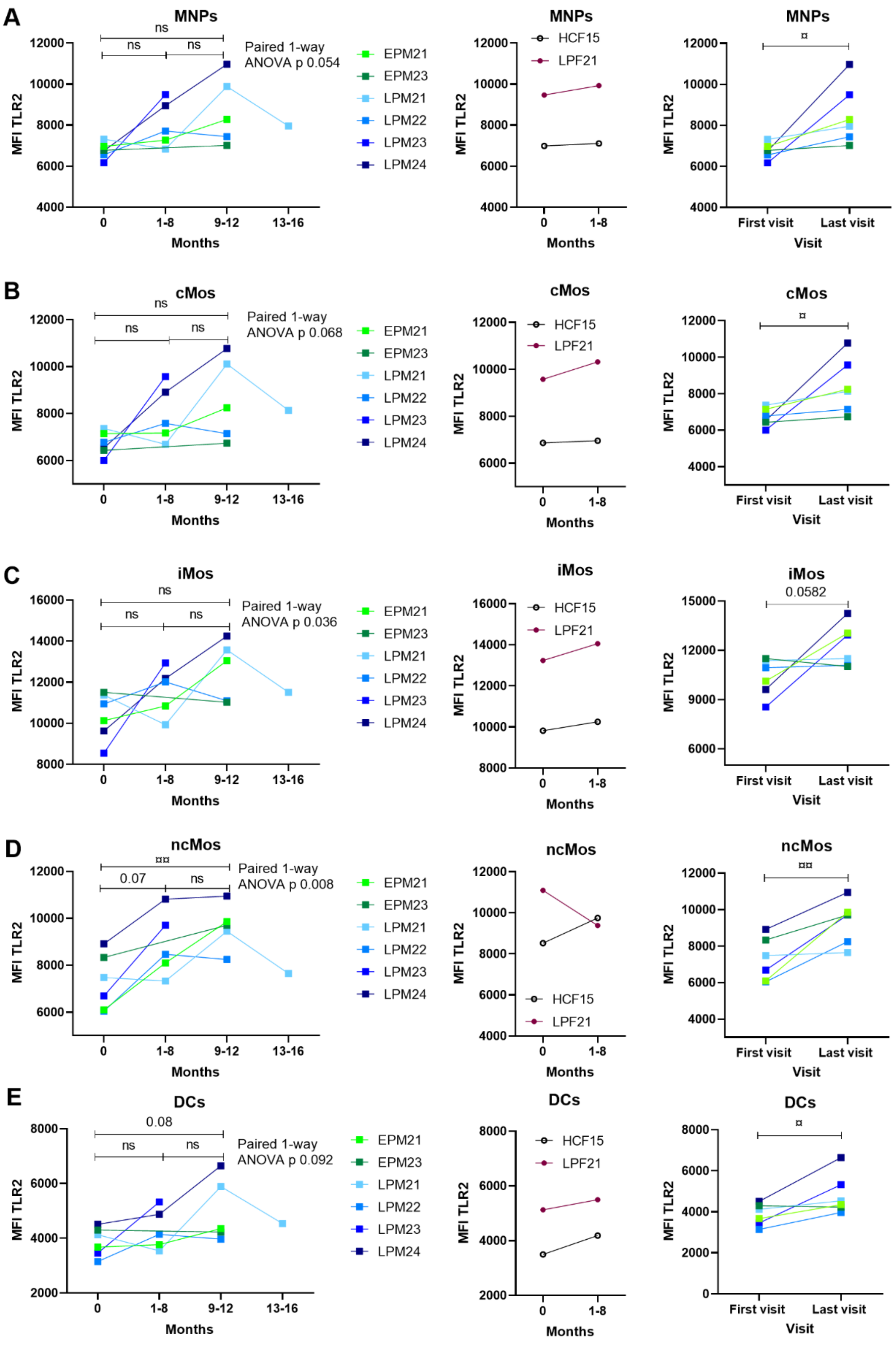
A
MNPs
ns
ns
ns
Paired 1-way ANOVA p 0.054
EPM21
EPM23
LPM21
LPM22
LPM23
LPM24
MFI TLR2
12000
10000
8000
6000
4000
0
1-8
9-12
13-16
Months
MNPs
HCF15
LPF21
MFI TLR2
Months
MNPs
¤
MFI TLR2
First visit
Last visit
Visit
B
cMos
ns
ns
ns
Paired 1-way ANOVA p 0.068
EPM21
EPM23
LPM21
LPM22
LPM23
LPM24
MFI TLR2
Months
cMos
HCF15
LPF21
MFI TLR2
Months
cMos
¤
MFI TLR2
First visit
Last visit
Visit
C
iMos
ns
ns
ns
Paired 1-way ANOVA p 0.036
EPM21
EPM23
LPM21
LPM22
LPM23
LPM24
MFI TLR2
16000
14000
12000
10000
8000
Months
iMos
HCF15
LPF21
MFI TLR2
Months
iMos
0.0582
MFI TLR2
15000
12000
9000
6000
First visit
Last visit
Visit
D
ncMos
¤¤
0.07
ns
Paired 1-way ANOVA p 0.008
EPM21
EPM23
LPM21
LPM22
LPM23
LPM24
MFI TLR2
Months
ncMos
HCF15
LPF21
MFI TLR2
Months
ncMos
¤¤
MFI TLR2
First visit
Last visit
Visit
E
DCs
0.08
ns
ns
Paired 1-way ANOVA p 0.092
EPM21
EPM23
LPM21
LPM22
LPM23
LPM24
MFI TLR2
8000
6000
4000
2000
Months
DCs
HCF15
LPF21
MFI TLR2
Months
DCs
¤
MFI TLR2
First visit
Last visit
Visit

**Supplementary Figure 6: Longitudinal comparison of TLR2 expression in males with PD**
Median fluorescents intensity (MFI) of TLR2 on **A)** all TLR2$^+$ mononuclear phagocytes (MNPs), and the separate subtypes: **B)** classical (cMos), **C)** intermediate (iMos), and **D)** non-classical monocytes (ncMos), as well as **E)** dendritic cells (DCs). Measured on sporadic Parkinson's disease (PD) patients with early (<5 years since diagnosis) or late (≥5 years since diagnosis) PD status and a single healthy control female (HCF) with multiple visit time points (*Cohort#1B*): Early PD male (EPM) (with progression to late (L)), late PD male (LPM) and female (LPF). **Left column:** graphs are showing monthly intervals of visits of all male patients: paired repeated measures mixed-effect model (REML) with Tukey's multiple comparisons; **middle column:** monthly intervals for LPF and HCF; **right column:** graphs are showing differences from first to last visit for all male patients: parametric paired t-test. ¤ $p<0.05$, ¤¤ $p<0.01$.
